# Seven chain adaptive immune receptor repertoire analysis in rheumatoid arthritis: association to disease and clinically relevant phenotypes

**DOI:** 10.1101/2021.11.26.21266347

**Authors:** Adrià Aterido, María López-Lasanta, Francisco Blanco, Antonio Juan-Mas, María Luz García-Vivar, Alba Erra, Carolina Pérez-García, Simón Ángel Sánchez-Fernández, Raimon Sanmartí, Antonio Fernández-Nebro, Mercedes Alperi-López, Jesús Tornero, Ana María Ortiz, Carlos Marras Fernández-Cid, Núria Palau, Wenjing Pan, Miranda Byrne-Steele, Dmytro Starenki, Daniel Weber, Ivan Rodriguez-Nunez, Jian Han, Richard M. Myers, Sara Marsal, Antonio Julià

## Abstract

Rheumatoid arthritis (RA) is an immune-mediated inflammatory disease characterized by a defective adaptive immune receptor repertoire (AIRR) that fails to distinguish self from non-self antigens. The AIRR is vast, encompassing four T cell receptor (TCR) and three B cell receptor (BCR) chains, each of which displays an extraordinary amino acid sequence variability in the antigen-binding site. How the concerted action of T and B cell clones is associated with the development and clinical evolution of immune-mediated diseases is still not known. Using a new immunosequencing technology that allows the unbiased amplification of the seven receptor chains, we conducted an in-depth quantitative analysis of the seven-receptor chain variability in RA. Compared to healthy controls, the AIRR in RA was found to be characterized by a lower BCR diversity, the depletion of highly similar BCR clones, an isotype-specific signature as well as a skewed IGL chain and gene segment usage. A predictor based on quantitative multi-chain AIRR information was able to accurately predict disease, including the elusive seronegative subset of RA patients. AIRR features of the seven immune receptor chains were also different between patients with distinct clinically relevant phenotypes. Incorporating HLA variation data, we were able to identify the TCR clones that are specifically associated with the main disease risk variants. The longitudinal analysis of the AIRR revealed that treatment with Tumor Necrosis Factor (TNF) inhibitors selectively restores the diversity of B cell clones in RA patients by reducing the frequency of clones with a similar biochemical profile. The biochemical properties of the TNFi-modulated clones were also found to differ between responders and non-responders, supporting a different antigenic reactivity in the B cell compartment of these two groups of RA patients. Our comprehensive analysis of the TCR and BCR repertoire reveals a complex T and B cell architecture in RA, and provides the basis for precision medicine strategies based on the highly informative features of the adaptive immune response.

## INTRODUCTION

Rheumatoid arthritis (RA) is a prevalent immune-mediated disease that is characterized by the chronic inflammation of the synovial membrane caused by inadequate activation of cells of the innate and adaptive immune systems^1^. Tumor necrosis factor (TNF) plays a central role in the inflammatory processes of RA, which can eventually lead to irreversible joint damage and bone erosion^2^. Although there is increasing knowledge on the immune cell types and states that participate in RA pathogenesis^3, 4^, little is known on how the immune repertoire -the collection of T and B cell receptor chains- is shaped by disease or how it is associated with phenotypes of interest like the response to TNF inhibition (TNFi) therapy.

The immune repertoire relies on T and B lymphocytes, two cell types that are responsible for regulating and executing the cell- and antibody-mediated adaptive immune responses^5^. The presence of RA-specific autoantibodies, IgM-rheumatoid factor (RF) and anti-citrullinated protein antibodies (ACPA) in a large percentage of patients is a hallmark of RA^6^. The pivotal role of B cells is also strongly supported by the efficacy of B cell depletion therapy in patients with RA^7, 8^. Similarly, blocking the activation of T cells has proven successful in controlling the disease activity^9^. The involvement of T cells in RA pathogenesis is also supported by the strong association between RA risk and genetic variation at the *HLA* class II locus, which determines antigen presentation to CD4+ T cells^10^. Therefore, the characterization of the immune repertoire of B and T cells is essential to better understand the misdirected adaptive immune response that is raised in RA.

In autoimmune diseases like RA, immune tolerance is compromised when T cell receptors (TCRs) and B cell receptors (BCRs) fail to distinguish self from non-self antigens^11^. Structurally, each TCR consists of two polypeptide chains (predominantly alpha-beta and gamma-delta pairs), and each BCR consists of four proteins, including two light chains (i.e., kappa or lambda) and two heavy chains. The latter determine the immunoglobulin (Ig) isotype, and therefore the functional properties of BCRs^12^. The collection of TCRs and BCRs present in an individual at a given moment constitutes the adaptive immune receptor repertoire (AIRR)^13^. The AIRR includes naive, but also effector as well as antigen-experienced memory cells that reflect the present and past of the individual’s immune responses, respectively^14^. The type and abundance of the seven receptor chains confer the functional diversity to the AIRR and, therefore, chain-wide analysis is fundamental to determine how the adaptive immune response is associated to disease and clinically relevant phenotypes.

The high diversity of T and B cell receptors that allows the recognition of an extremely broad space of potential antigens is one of the most important hallmarks of the AIRR^15^. This high diversity emerges as a consequence of the genomic rearrangement between the variable (V), diversity (D) and joining (J) AIRR gene segments, along with the addition and removal of random nucleotides^16, 17^. The resulting variability in the amino acid sequence of the complementarity-determining region 3 (CDR3) that spans the V(D)J junction is key to determine antigen binding and specificity^18, 19^. In BCRs, antigen-binding affinity is further diversified through the somatic hypermutation of V gene segments^20^. Additionally, BCR can undergo class switching during B cell differentiation in the germinal center, conferring distinct immunological roles to the antibody^21, 22^. As recently described in other autoimmune diseases^23^, the isotype signatures specific to B cells could provide new insights into the misdirected autoimmune response and the generation of different pathological features in RA.

Recent advances in high-throughput sequencing technologies are allowing the quantitative analysis of the immune repertoire at unprecedented depth and resolution^24^. AIRR immunosequencing (AIRR-seq) is proving successful for disease detection and monitoring of treatment response in cancer and infectious diseases^25–27^. To date, however, only a few studies analyzing the peripheral repertoire have been performed in RA^28, 29^. These analyses have been restricted to one type of receptor, limiting the global understanding of the complexity and interconnectedness of the adaptive response. Very recently, a new AIRR-seq technology that performs an unbiased analysis of the seven receptor chains in a single assay has been developed^30^. Coupling this technology with new methodological approaches for AIRR-seq analysis that allow to distinguish true biological signals from stochastic processes^31^, the longitudinal analysis of patients can reveal pivotal insights to better understand how the immune repertoire is altered by disease and by treatment.

In our work, we have characterized the immunogenomic profile of RA. Using case-control, case-case and longitudinal study designs, we have comprehensively analyzed the chain-wide AIRR variability association with disease and with clinically-relevant phenotypes, including the response to TNFi therapy. The present study demonstrates the potential of AIRR genomic information for the development of new immune receptor-based precision medicine strategies in autoimmunity.

## RESULTS

### Quantitative characterization of the AIRR from RA patients and healthy controls for the seven receptor chains

To conduct a comprehensive AIRR-seq analysis of the seven receptor chains in RA, we immunosequenced whole blood samples from 104 healthy controls and 86 RA patients. All RA patients were analyzed at the beginning of TNFi therapy and 77 of them were also immunosequenced after 12 weeks of treatment (**Table S1**). For each sample, an unbiased amplification of the TCR-alpha (TRA), TCR-beta (TRB), TCR-delta (TRD), TCR-gamma (TRG), BCR-Ig heavy (IGH), BCR-Ig lambda (IGL) and BCR-Ig kappa (IGK) chains was performed in a single assay (**Fig. 1a**). The entire chain-wide sequencing of our study population yielded an average of >5 and >6.5 million assembled and demultiplexed reads per sample, respectively (**Table S2**). These immune receptor reads were found to map to >28.5 million unique molecular identifiers (UMI) including >3.5 million UMIs for each IGH, IGL and IGK chains, >6.2 million UMIs for each TRA and TRB chains as well as >150,000 UMIs for each of the less abundant TRD and TRG chains (**Table S2**). Altogether, chain UMIs provided quantitative information for >8.2 million unique clones that were available for analysis (**Table S2**). In order to identify potential confounding factors, we first evaluated the impact of technical and epidemiological variables over the AIRR properties. We found that sequencing depth had a significant impact on multiple AIRR measures (**Fig. S1**), particularly on the TRB diversity. We also detected a significant association between the patient’s gender and IgD isotype levels (**Fig. S1**). To control for these confounders, all subsequent AIRR analyses were adjusted for the sequencing depth as well as for sex and age.

**FIGURE 1.**
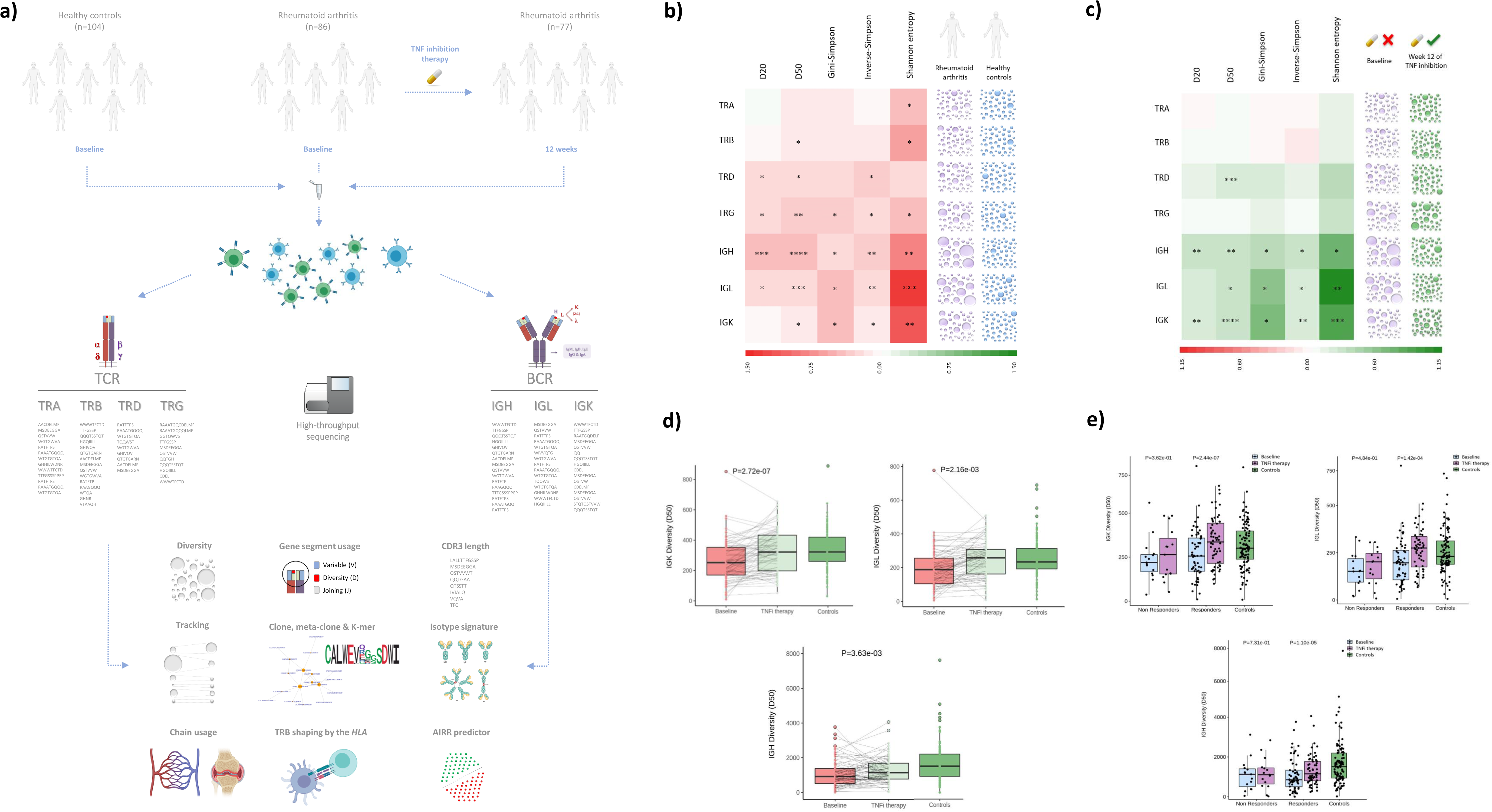
Overview of the study design and chain-wide characterization of the AIRR diversity in RA. a) Overview of the study design. b) Chain-wide characterization of the AIRR diversity in rheumatoid arthritis compared to controls. This includes the effect that disease have on five complementary diversity measures (i.e., D20, D20, Gini-Simpson, Inverse-Simpson and Shannon entropy) for each of the seven immune receptor chains. The heatmap is colored according to the effect size of the association, ranging from red (i.e., diversity reduction) to green (i.e., diversity increase). The graphical explanation of the association results at the chain level is shown on the right side. Abbreviations: *, P<0.05; **, P<5.00e-03; ***, P<5.00e-04; ****, P<5.00e-05. c) Chain-wide impact of TNF inhibition therapy on the AIRR diversity in rheumatoid arthritis. The parameters used for the graphical representation are described in b. d) Overall impact of TNFi therapy on the AIRR diversity measured by the D50 index in the three BCR chains. Abbreviations: P, p-value. e) Impact of TNFi therapy on the AIRR diversity measured by the D50 index in the three BCR chains stratified by clinical response to TNF inhibition. Abbreviations: P, p-value.

### The low diversity that characterizes the AIRR of RA patients is mainly driven by the expansion of BCR clones and is restored by TNFi therapy

To investigate whether the immune repertoire diversity is altered in RA, we compared the clone diversity between patients and controls. We found that clone diversity in RA is significantly reduced in all seven chains compared to healthy individuals (**Fig. 1b**). The most significant diversity reduction occurred in the B cell compartment, including IGH (*P_D50_*=4.22e-05), IGL (*P_D50_*=3.30e-04) and IGK (*P_D50_*=9.13e-03). The observed differences were independent of the type of diversity index used (**Table S3**).

At the clinical level (**Fig. S2, Table S4**), the most consistent signal across diversity measures was found to be a significant increase of clone diversity of the IGK chain in ACPA-positive patients in comparison to ACPA-negative patients (*P_D50_*=9.12e-03). We also detected a significant association between the clinical response to TNFi therapy and the diversity levels of TRG (*P_Inverse-Simpson_*=2.70e-02). In particular, responder patients showed a higher diversity of TRG clones than non-responders. TRD chain clones also showed a reduced diversity in both ACPA- (*P_D50_*=2.89e-02) and RF-positive patients (*P_Shannon_*=4.62e-02). These results indicate that clinically-relevant phenotypes in RA also are associated with AIRR features.

The longitudinal analysis showed that TNFi therapy has a strong impact on the BCR repertoire (**Fig. 1c**). Diversity in IGK (*P_D50_*=2.72e-07), IGH (*P_D50_*=3.63e-03) and IGL (*P_D50_*=2.16e-03) chains was found to increase after three months of systemic drug therapy (**Fig. 1d, Table S5**). These findings show that blocking of TNF contributes to restore the BCR clone diversity towards that observed in healthy individuals (**Fig. 1d**). Stratifying the longitudinal analysis by clinical response, the TNFi effect on the diversity of BCR chains was found to occur only in the responder group of patients (*P_D50_*<1.43e-04, **Fig. 1e**). Like in the case-control analysis, the longitudinal results were significant and independent of the immune repertoire diversity measure used (**Table S6**).

### Identification and characterization of contracted and expanded clones after TNFi therapy

The finding that BCR diversity is restored by TNFi therapy suggests that the contraction of B cell clones is a downstream effect of the drug’s mechanism of action. To identify the BCR clones that are contracted or expanded during the 12 weeks of therapy, we used a Bayesian approach that accounts for the specific statistical features of longitudinal AIRR-seq data^31^. Since B cell activation is mostly T-cell dependent, we also analyzed the clonal expansion of TCR clones. Using this approach, we identified n=42,488 and n=55,195 clones from both immune cell receptors that are significantly contracted and expanded after TNFi therapy, respectively (**Table S7**). Consistent with the findings obtained in the clone diversity analysis, the number of contracted clones was found to be significantly higher in the BCR (*N*=41,863 clones, 2.50%) than in TCR (*N*=625 clones, 0.016%) chains (*P*<2.20e-16, **Fig. 2a**). Clonal expansion was found to be also more marked in the B cell compartment (*N*=54,433 BCR clones, 1.86%) than in T cell compartment (*N*=762 TCR clones, 0.020%, *P*<2.20e-16, **Fig. 2a**). The net effect of both processes might suggest a predominant clonal expansion induced by TNF blocking. However, comparing the magnitude of the clonal frequency variation induced by TNFi therapy, we detected that it is significantly higher in contracted clones from IGH, IGL, IGK, TRA, TRB and TRG chains (*P*<0.05, **Fig. 2b**). The strongest differences were detected in the BCR chains (*P*<4.40e-14, **Fig. 2b**), supporting the findings obtained in the clone diversity analysis.

**FIGURE 2.**
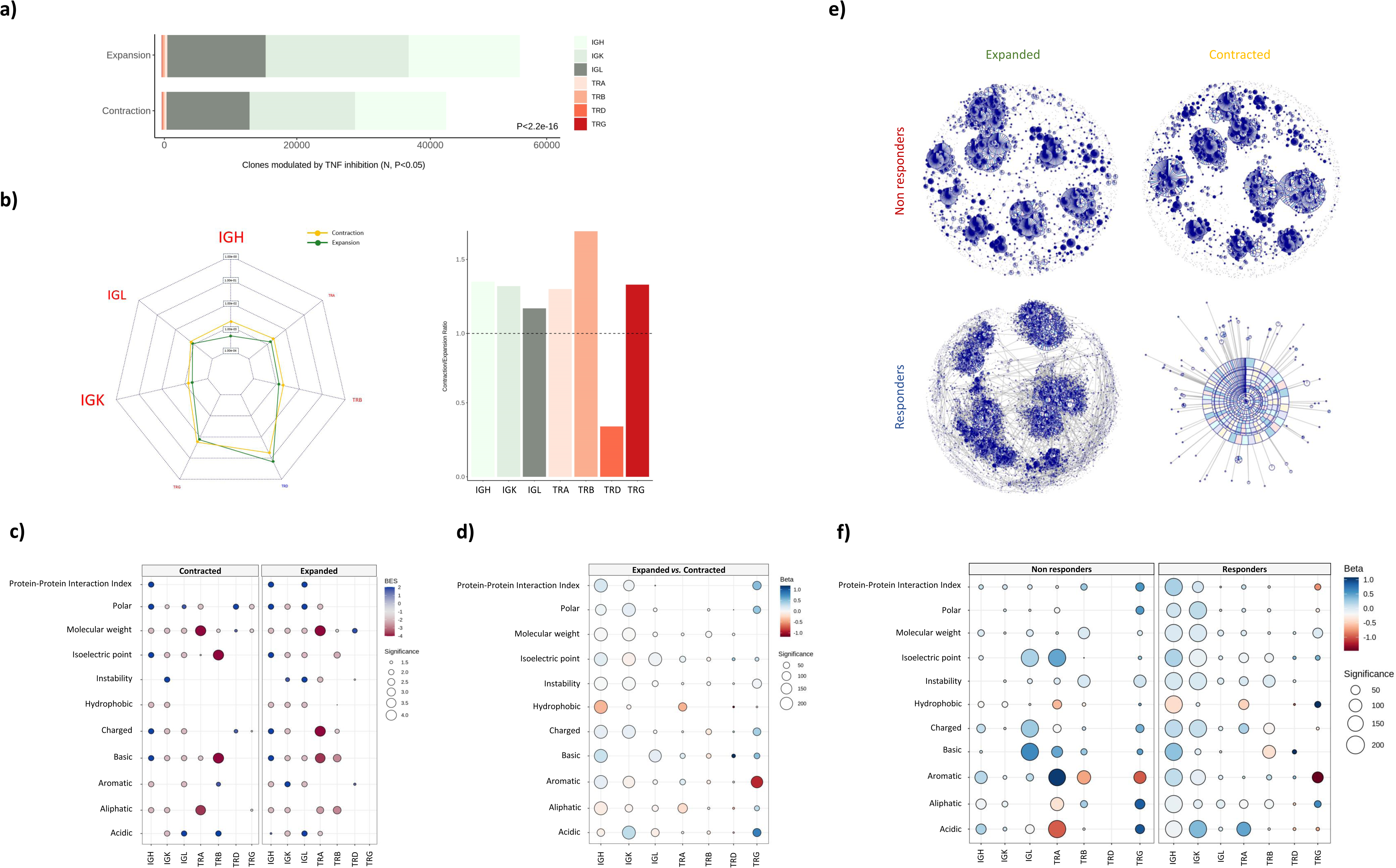
Identification and characterization of contracted and expanded clones after TNF inhibition. a) Number of clones that are significantly contracted and expanded after 12 weeks of TNF inhibition (P<0.05). Abbreviations: N, number of clones; P, p-value. b) In the left side, the average clonal frequency variation detected in contracted and expanded clones is shown at the chain level. The scale of each radar plot break is independent from the others. The size of the chain name is proportional to the statistical significance of the difference of clonal frequency variation between contracted and expanded clones. Chains showing a significantly different clonal frequency variation between contracted and expanded clones are colored in red (P<0.05). In the right side, the ratio between the average clonal frequency variation detected in contracted and expanded clones is shown. c). Biochemical profile of significantly contracted and expanded clones after TNFi therapy. For each chain, the enrichment of the indicated group of clones in each biochemical feature is represented by a circle. The circle is colored according to the degree of biochemical enrichment (i.e., blue) or depletion (i.e., red) compared to a random repertoire from the same chain. The size of the circle is proportional to the statistical significance of the enrichment in each biochemical feature. Only significant differences are represented as circles (P<0.05). Abbreviations: BES, Biochemical Enrichment Score. d) Differential biochemical profile resulting from directly contrasting expanded and contracted clones after TNFi therapy. For each chain, the association of clonal contraction/expansion with the indicated biochemical property is represented by a circle. The circle is colored according to the effect direction, ranging from red (i.e., the biochemical feature is lower in contracted clones) to blue (i.e., the biochemical feature is higher in expanded clones). The size of the circle is proportional to the statistical significance of the association. Non-significant differences are hidden (P>0.05). e) CDR3 amino acid sequence similarity networks for expanded and contracted IGL clones stratified by clinical response to TNFi therapy. Clones are represented as nodes and edges connect clones differing by at most one amino acid in the CDR3 sequence. The node size is proportional to the number of similarity connections with other clones. Each node is divided into as many fractions as individuals have the clone. f) Differential biochemical profile resulting from directly contrasting expanded and contracted clones within each group of responder and non-responder patients to TNFi therapy. The details of the graphical representation are described in d.

Based on the previous results, we hypothesized that the clones that are affected by TNFi therapy might be reacting to the same antigens. To test this, we evaluated the level of sequence similarity and its potential functional role. We found that contracted clones from the seven receptor chains have a higher CDR3 amino acid sequence similarity than expected by chance (*P*<1e-04, **Table S8**), a property that was also detected among expanded clones (**Table S8**). This similarity was also associated to the presence of distinctive biochemical features between the two types of dynamic changes (**Fig. 2c**). While many of the biochemical properties were shared between the contracted and expanded clones (e.g., low molecular weight in clones from the three BCR chains; P<1e-04), there were also contraction- and expansion-specific biochemical features like low charge (P<1e-04) and high protein-protein interaction index detected in IGL clones (P<1e-04), respectively. Biochemical differences were even more accentuated when directly contrasting the abundance of expanded and contracted clones. In this analysis, IGH clones that are contracted by TNFi therapy are significantly much less hydrophobic than expanded clones (P<1.00e-220, **Fig. 2d**). The restriction of the therapeutic effect to clones with a specific biochemical profile, particularly in the IGH and IGK BCR chains, suggests that the modulation of the immune repertoire by TNFi is highly antigen-specific.

We further investigated the properties of the clones modulated by TNFi therapy according to the two levels of clinical efficacy. In both responder and non-responder patients, the similarity of the CDR3 amino acid sequences of the contracted and expanded clones was found to be higher than expected by chance in all receptor chains (*P*<1e-04, **Fig. 2e, Table S9**). By directly contrasting the amino acid properties between contracted and expanded clones in each group of patients, the biochemical profile of the clones modulated by TNF inhibition was found to differ between responder and non-responders (**Fig. 2f**). Responders to TNFi therapy showed the strongest biochemical differences in IGH and IGK clones (P<7.45e-10). In non-responders the differences in these two chains were moderate and, instead, marked differences occurred in the biochemical profile of clones from IGL and TRA chains (P<5.95e-05). These results suggest that patients who will respond to therapy have a different antigenic reactivity in the B cell compartment from those that will not show clinical improvement, and that this reactivity is sensitive to systemic TNF levels.

### Low IGL usage in blood of RA patients suggests increased migration rate of IGL-harboring B cells to the synovial tissue

To explore whether RA affects the abundance of each receptor chain, we compared the chain usage between cases and controls. Among all seven chains, we found that IGL is less abundant in the blood of RA patients than in healthy subjects (P=2.50e-03, **Fig. 3a**). This reduction was not observed in the IGK chain and, consequently, the small chain immunoglobulin ratio IGK/IGL was significantly altered by disease (1.70 detected in the healthy cohort *vs.* 2.14 in RA, P=1.03e-03). Based on this observation and the importance of B cells in RA pathology, we hypothesized that the circulating levels of IGL detected in RA patients could be explained by an increased migration of IGL-harboring B cells from blood to synovium. To investigate this, we analyzed the immune repertoire in paired blood and synovial samples using an available bulk RNA-seq dataset from RA patients^32^. In line with our hypothesis, we found that IGL usage is significantly higher in the RA synovium compared to peripheral blood (P=4.90e-09, **Fig. 3b**).

**FIGURE 3.**
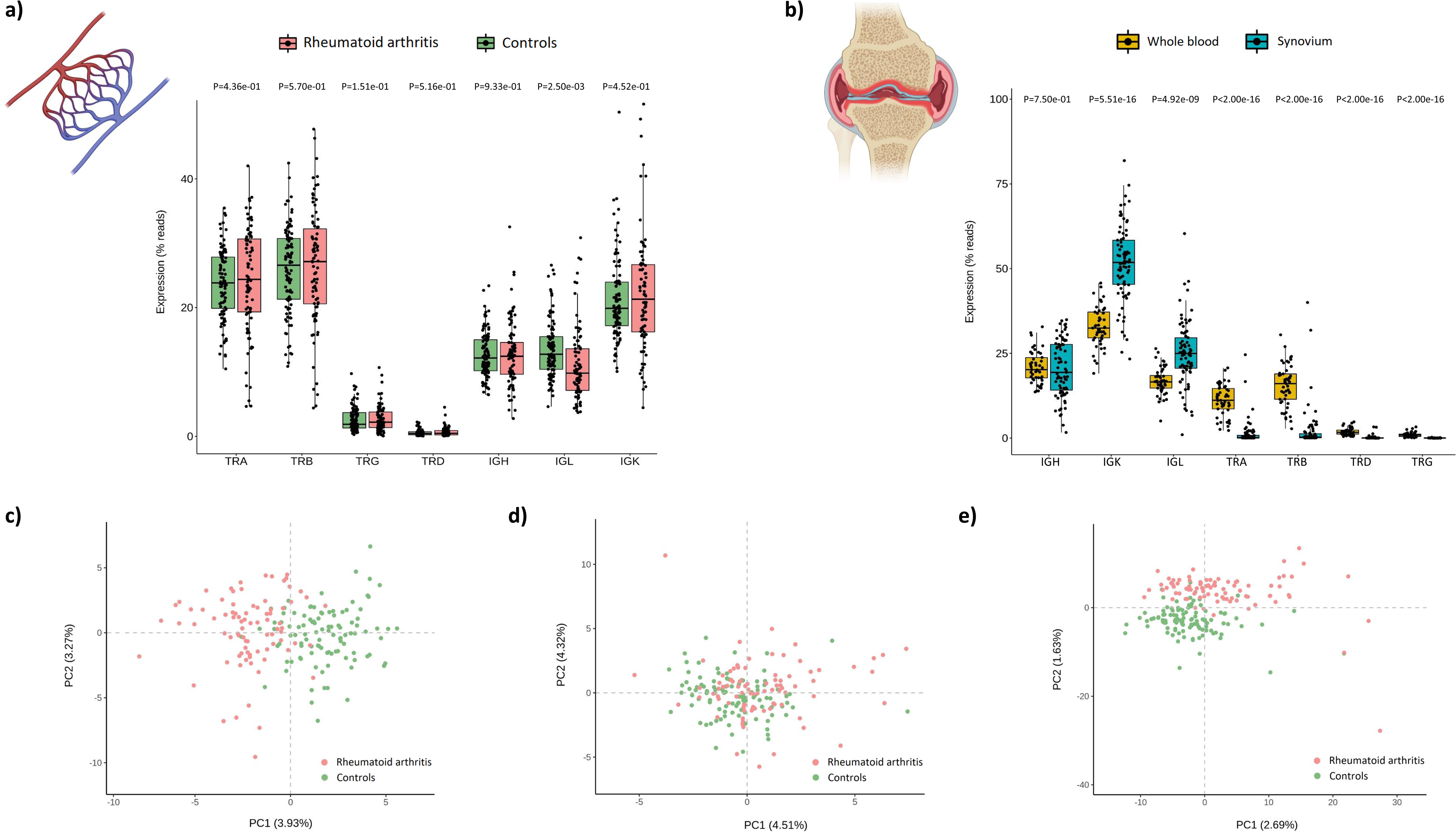
Chain and gene segment usage profiles of rheumatoid arthritis patients. a) Differences in the whole blood usage of each immune receptor chain between rheumatoid arthritis and healthy individuals. b) Differences in the usage of each immune receptor chain between whole blood and synovium samples from patients with rheumatoid arthritis. c) First two principal components of the V gene segment usage (N=298 variables from the seven immune receptor chains) in rheumatoid arthritis and controls. d) First two principal components of the J gene segment usage (N=100 variables from the seven immune receptor chains) in rheumatoid arthritis and controls. e) First two principal components of the usage of V-J segments pairs (N=4,054 variables from the seven immune receptor chains) in rheumatoid arthritis and controls. Abbreviations: P, p-value; PC, principal component.

Unlike the repertoire diversity, chain usage was not significantly associated with clinical phenotypes. Only negative trends between TRA usage and ACPA positivity and between TRD usage and disease activity were found (**Fig. S3**). Also, the longitudinal analysis showed that neither TCR or BCR chain usage is significantly altered by TNFi therapy (**Table S10**).

### RA patients are characterized by a skewed usage of V and J gene segments

There is growing evidence that alterations in the patient’s AIRR can be reflected in a biased usage of V and J gene segments^33^. In line with this, we found that the usage profile of V segments and V-J pairs is very different between RA and control subjects (**Fig. 3c-e**). When performing the comparative analysis stratified by segment type, the usage of 34 V segments, 12 J segments and 127 V-J pairs was found to be significantly different between RA and control subjects (*FDR*<0.05, **Table S11**).

Chain usage was more moderately associated to clinical phenotypes and the response to therapy. A total of 28, 34 and 234 nominal associations were identified with the usage of V segments, J segments and V-J pairs, respectively (*P*<0.05, **Table S12**). The top association signal was detected between disease activity and TRAV23DV6-TRAJ22 segment pair (*P*=1.12e-03). The combination of IGKV2-28 and IGKJ1 segments showed the strongest association with TNFi response (*P*=1.98e-03), and TRDV1-TRDJ1 was found to be the most strongly associated segment pair with both the RF and ACPA clinical phenotypes (*P*=3.45e-03). In the longitudinal analysis (**Table S13**), TNFi treatment was found to moderately regulate the usage of two V segments from IGH (*P*=1.39e-02) and IGK (*P*=3.34e-02) as well as 10 V-J segment pairs (*P*<0.05), indicating that TNFi therapy has a little impact on the gene usage profile in RA patients.

### Identification and characterization of RA-associated clones, meta-clones and k-mers

Analysis at the single-clone level identified a total of 20 IGL and 40 IGK clones with a significantly lower abundance in RA patients compared to healthy controls (*FDR*<0.05; **Fig. 4a-b, Table S14, Fig. S4**). The similarity of the CDR3 amino acid sequence across these clones was found to be significantly higher than expected by chance (*P<1.00e-09*, **Fig. 4c, Table S15**). This result suggests that these clones reduced in RA have reactivity to a common antigen^34^. By screening the Pan Immune Repertoire Database of T and B cell receptors targeting known antigens^35^, we found that none of the RA-associated IGL and IGK clones recognize any of the antigens that have been discovered so far. Compared to the biochemical properties of a random repertoire, we found that the IGL clones downregulated in RA are more acidic and have a lower isoelectric point than expected by chance (*P*<0.05, **Fig. 4d**). IGK clones were found to be more hydrophobic and aliphatic, but less polar (*P*<0.05) than expected by chance (**Fig. 4d**). No clones passed the multiple testing threshold in the clone-wide association analysis with clinical phenotypes (**Table S16**). However, analyzing the set of clones associated with RA, we found that the levels of IGK chain clone CMQALQTPPYTF at baseline are increased in non-responders to TNFi therapy compared to responders (*FDR*=1.25e-02, **Table S17**).

**FIGURE 4.**
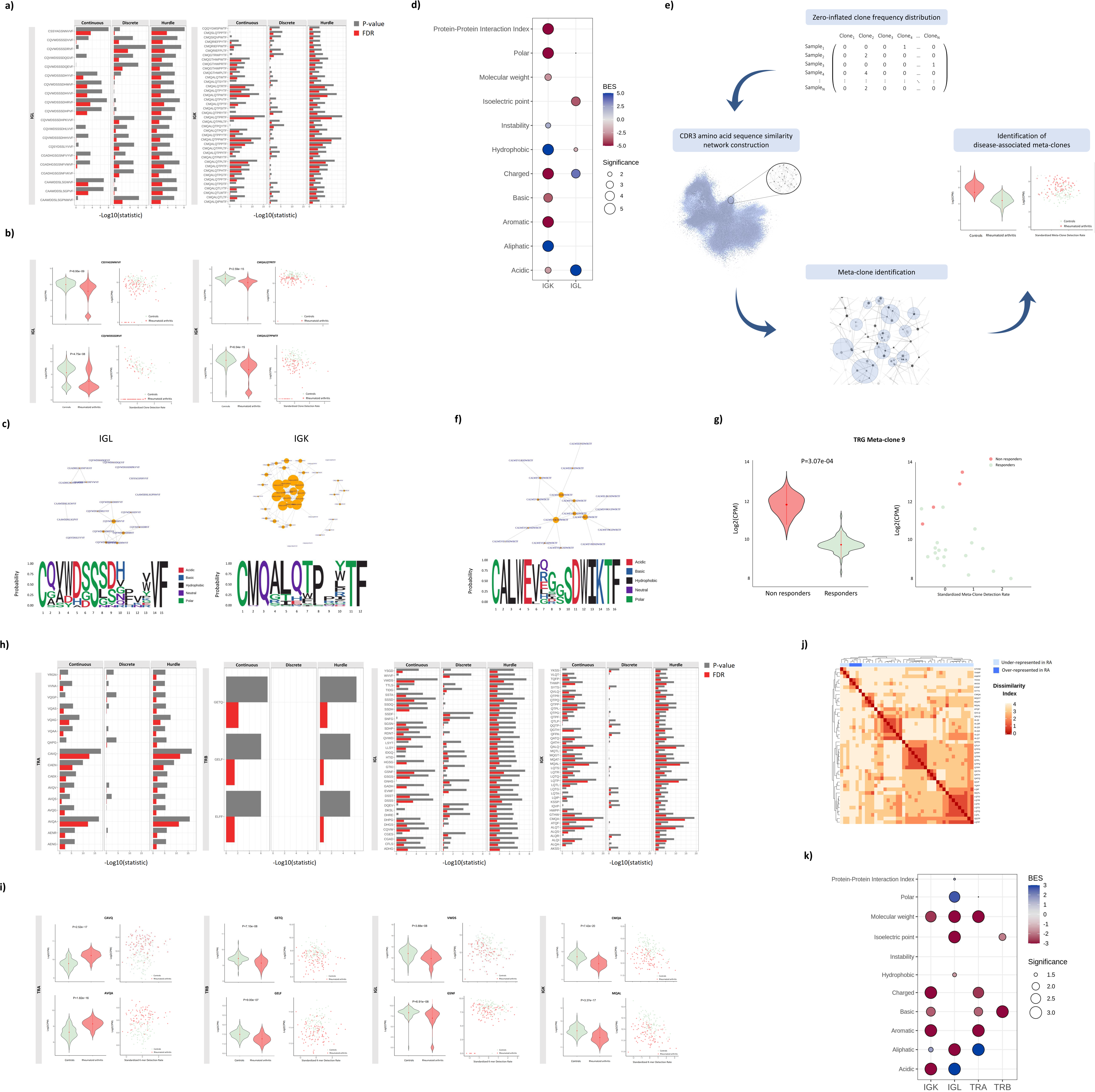
Identification and characterization of clones, meta-clones and k-mers associated with rheumatoid arthritis. a) CDR3 amino acid sequences of the clones showing a significantly different abundance in rheumatoid arthritis compared to healthy controls (FDR<0.05). Abbreviations: FDR, false discovery rate. b) On the left side, the expression of the two clones showing the strongest association with rheumatoid arthritis is shown. On the right side, the clone expression is plotted against the standardized clone detection rate, indicating that the associations detected are not influenced by the sampling bias. Abbreviations: CPM, counts per million; P, p-value. c) CDR3 amino acid sequence similarity networks and amino acid motif of the IGL and IGK clones associated with rheumatoid arthritis. In the similarity networks, each clone is represented as a node and edges connect clones differing by at most one amino acid in the CDR3 sequence. The node size is proportional to its number of similarity connections with other clones. The p-value shown for each chain indicates the probability of observing the indicated similarity by chance. Abbreviations: P, p-value d) Biochemical profile of the clones significantly associated with rheumatoid arthritis. For each chain, the enrichment of the associated clones in each biochemical feature is represented by a circle. The circle is colored according to the degree of biochemical enrichment (i.e., blue) or depletion (i.e., red) compared to a random repertoire from the same chain. The size of the circle is proportional to the statistical significance of the enrichment in each biochemical feature. Abbreviations: BES, Biochemical Enrichment Score. e) Overview of the meta-clone analytical strategy used here to identify groups of highly similar clones associated with disease conditions. Abbreviations: CPM, counts per million. f) CDR3 amino acid sequence similarity network and amino acid motif of the meta-clone 9 from TRG chain that showed a significant association with the clinical response to TNFi therapy. The network parameters are described in c. g) On the left side, expression levels of the meta-clone 9 from TRG in responder and non-responder patients to TNFi therapy. On the right side, the expression of meta-clone 9 is plotted against the standardized meta-clone detection rate. Abbreviations: CPM, counts per million; P, p-value. h) K-mers showing a significantly different abundance in rheumatoid arthritis compared to healthy controls (FDR<0.05). Abbreviations: FDR, false discovery rate. i) On the left side, the expression level of the two k-mers showing the strongest association with rheumatoid arthritis is shown. On the right side, the k-mer expression is plotted against the standardized k-mer detection rate. Abbreviations: CPM, counts per million; P, p-value. j) Pairwise dissimilarity matrix among IGK k-mers significantly associated with RA (FDR<0.05). K-mers over- and under-represented in rheumatoid arthritis are represented in dark and light blue, respectively. The dissimilarity index was computed using the Levenshtein distance measure so that the higher the Levenshtein distance, the higher the dissimilarity index between two k-mer sequences. Abbreviations: RA, rheumatoid arthritis. k) Biochemical profile of the k-mers significantly associated with rheumatoid arthritis. The graphical parameters are described in d.

To identify groups of highly similar clones associated with RA and clinical phenotypes we developed the meta-clone method (**Fig. 4e**). Using this approach, we identified 3 IGL and 1 IGK meta-clones associated with RA (*FDR*<0.05, **Table S18**). Unlike the clone-level analysis, the meta-clone approach was able to identify a significant association with RA phenotypes (**Table S19**). Before initiating TNFi therapy, a meta-clone from the TRG chain was found to be significantly increased in non-responders compared to responder patients (meta-clone 9 with *FDR*<0.05, **Fig. 4f-g**).

Since it is only a fraction of the amino acids of the CDR3 sequence that directly interacts with the antigen, we also sought to investigate the difference in abundance of smaller amino acid motifs or k-mers in RA clones compared to healthy subjects. Consistent with the results in the single-clone and meta-clone association analyses, we found n=43 IGK and n=38 IGL k-mers that are differentially represented in RA compared to controls (*FDR*<0.05, **Table S20,** **Fig. 4h**). In addition to these associations, we also identified 16 k-mers from TRA and 3 k-mers from TRB significantly associated with RA (*FDR*<0.05, **Table S20,** **Fig. 4h**). From the total of 100 RA-associated BCR and TCR k-mers, 22 and 78 k-mers were found to be over and under-represented in RA, respectively (**Fig. 4i, Fig. S5**). All RA-associated k-mers displayed a high sequence similarity (*P<1.00e-04*, **Table S21**). From these, only IGK k-mers over-represented in RA were found to cluster together according to their sequence similarity (**Fig. 4j, Fig. S6**). RA-associated k-mers from different chains were found to have a significantly different biochemical profile (**Fig. 4k**). In the case-case analysis, k-mers in IGK (n=3) and IGL (n=16) chains were also significantly associated with clinical phenotypes (**Table S22, Table S23**). The strongest k-mer associations were detected with TNFi response and disease activity (*FDR*<0.05, **Table S22**). In particular, the DHRE k-mer was found to be detected only in responders to TNFi therapy (*FDR*=1.21e-02) and the WVVF k-mer showed a significantly higher abundance in patients with low disease activity compared to high disease activity patients *FDR*=2.16e-02).

### *HLA* alleles associated with RA risk mediate the antigen presentation to expanded and disease-specific TRB clones

Antigenic peptide residues presented by HLA proteins are directly in contact with amino acids spanning the CDR3 sequences from the TRB chain. Based on this antigen presentation mechanism, we sought to find how HLA variation shapes TRB clone expansion. Using the grouping lymphocyte interactions by paratope hotspots (GLIPH) algorithm ^36^, we identified a total of 126 and 504 clusters of TRB clones that are significantly enriched in the immune repertoire of RA patients compared to a naive repertoire of CD4+ and CD8+ T cells, respectively (*FDR*<0.05, **Table S24**). The little overlap observed between significant clusters from CD8+ and CD4+ T cells (i.e., 2.06%) indicates a marked cell-type specificity for these groups of TRB clones. From the total of 630 clusters of TRB clones that are over-represented in RA, 417 were found to be also significantly enriched in the immune repertoire of healthy controls (*FDR*<0.05, **Table S24**). These findings suggest that the remaining 213 clusters of TRB clones are RA-specific (n=56 CD4+ T cell clusters; n=157 CD8+ T cell clusters). Amino acid motif analysis of each cluster revealed seven RA-specific motifs that have a significantly different abundance between RA and control individuals (*P*<0.05, **Table S20**).

We further investigated whether the antigen presentation to the RA-specific clusters of TRB clones is modulated by *HLA* class II alleles. In this analysis, we found that the *HLA* class II haplotypes are likely to restrict the antigenic peptide presentation to 65 out of the 213 RA-specific clusters (*P*<0.05, n=16 CD4+ T cell clusters; n=49 CD8+ T cell clusters, **Fig. 5**). The strongest signal from CD4+ T cells was detected between the *HLA-DRB1*15* allele and the TRB cluster (QDFA amino acid motif, *P*=4.20e-03, **Fig. 5**). Association analysis of *HLA* haplotypes in a large case-control cohort of the same population (n=1,191 RA patients and 1,558 healthy controls), corroborated that *HLA-DRB1*15* is also the *HLA* variant that is most strongly associated with RA risk in Spain (*P*=8.86e-11, **Table S25**). Analysis of the QDFA motif in RA phenotypes showed that it is also significantly increased in ACPA-negative patients compared to ACPA-positive patients (*P*<0.05, **Table S23**). These findings demonstrate the functional impact of genetic risk variation on the shaping of the immune repertoire in RA.

**FIGURE 5.**
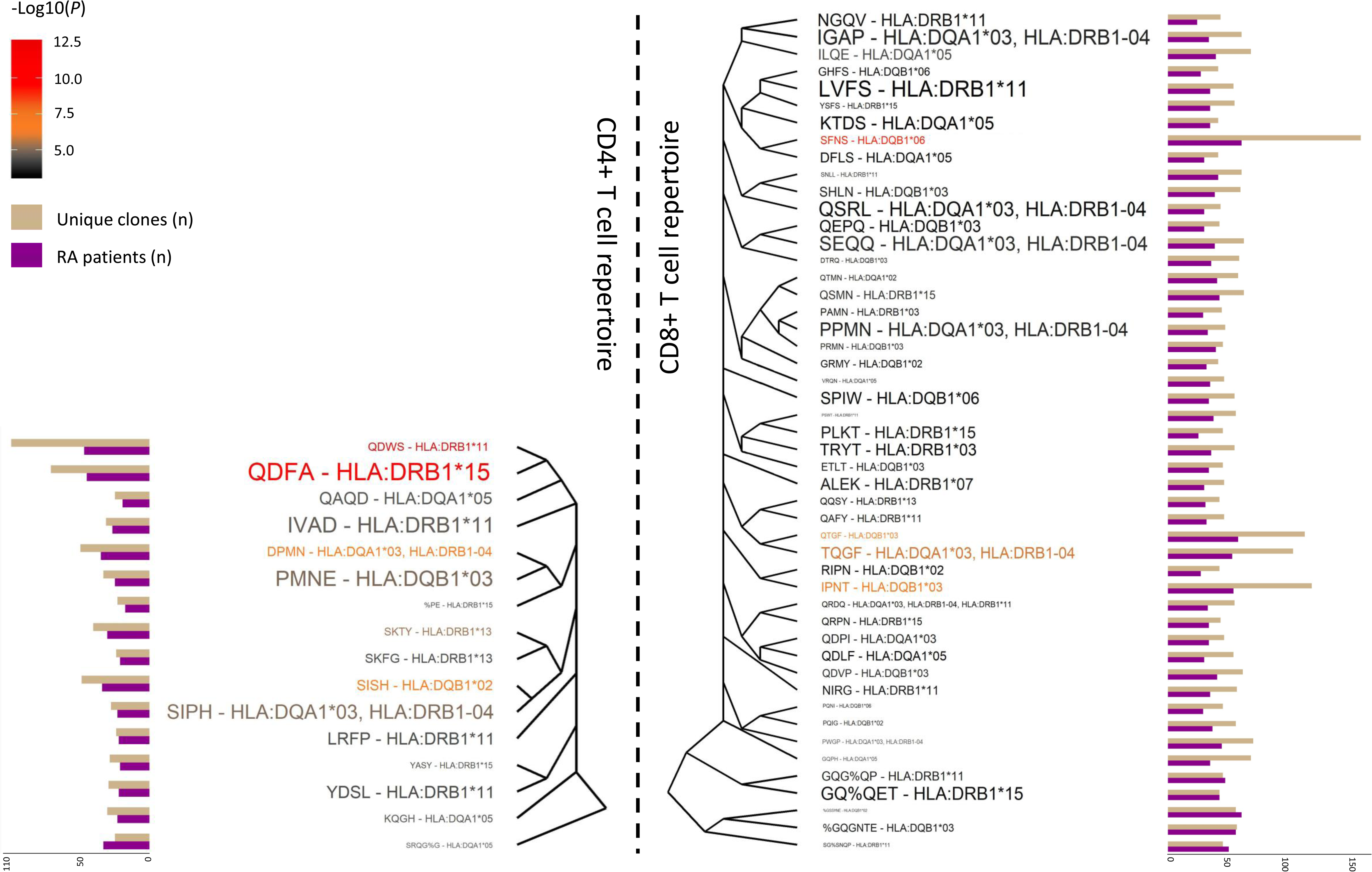
Clusters of TRB clones that are specific to rheumatoid arthritis and associated with HLA class II alleles. Clusters of TRB clones that are associated with HLA class II alleles and specifically enriched in the immune repertoire of rheumatoid arthritis patients compared to a naive repertoire of CD4+ (left side) and CD8+ T cells (right side). The most representative amino acid motif of those clones that are clustered together is used for the cluster naming. Clusters of TRB clones are sorted according to the Levenshtein distance among amino acid motifs. The number of unique clones belonging to each cluster and the number of patients having the indicated cluster are used for the cluster characterization. Each motif-allele pair is colored in accordance with the statistical significance of the motif enrichment in rheumatoid arthritis. The size of each association is proportional to the statistical significance of the genetic association between the indicated HLA allele and amino acid motif. The name of each amino acid motif is based on the nomenclature used by the GLIPH algorithm. Abbreviations: N, sample size, P, p-value; RA, rheumatoid arthritis.

### The length of the CDR3 amino acid sequence is associated with disease, clinical phenotypes and anti-TNF response

The length of the CDR3 amino acid sequence has been previously associated with polyreactivity and increased risk for self-recognition and autoimmunity^37^. In the present study, we found a highly significant enrichment of long TRD in RA compared to healthy individuals (*P*=7.91e-202, **Fig. S7**). This signal was mainly driven by the over-representation of TRD clones with more than 20-23 amino acids of length (**Fig. S7**). Clone length was also found to be significantly associated with clinical phenotypes (**Fig. S7).** For example, the length of TRD clones was found to be significantly different between ACPA-positive and ACPA-negative patients (*P*=6.69e-260), and between high and low disease activity patients (*P*=2.24e-08). The longitudinal analysis revealed that TNFi therapy alters the clone length distribution within the IGH repertoire (**Fig. S7**). Stratifying the analysis by clinical response, we found that the proportion of IGH clones with less than 18-20 amino acids is reduced in responders towards the levels observed in controls. An opposite effect was detected in non-responder patients to TNFi therapy (**Fig. S7**).

### RA patients display an isotype-specific signature

To further characterize properties of the BCR repertoire in RA, we analyzed the isotype information contained within the constant region of the IGH chain. This region interacts with effector molecules and cell types and therefore determines the functional role of the immunoglobulin. In this analysis (**Fig. 6a**), the levels of IgD isotype were found to be significantly lower in RA compared to controls (*P*=4.45e-04). Conversely, RA patients showed higher levels of IgG than healthy subjects (*P*=5.30e-03). None of the five isotypes showed a significant association with RA clinical phenotypes (**Table S26**).

**FIGURE 6.**
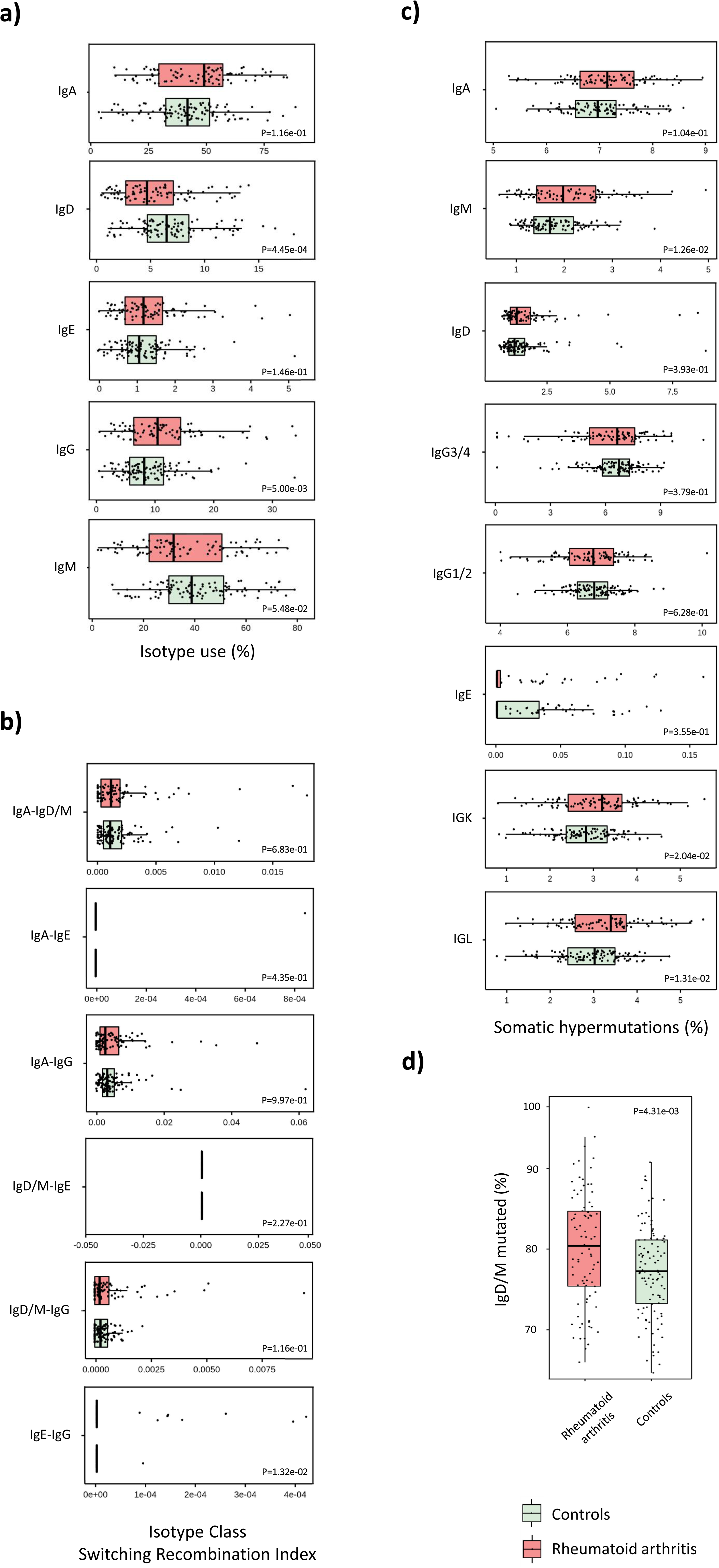
Characterization of the isotype signature underlying rheumatoid arthritis. Comparison of the a) isotype usage, b) isotype class switching index, c) somatic hypermutation profile and d) percentage of IgD/M mutated between patients with rheumatoid arthritis and healthy controls. Abbreviations: P, p-value.

After a B cell is activated by an antigen in an immune response, BCRs acquire distinct functional roles through isotype class switching. We next compared the degree of isotype class switching between RA and healthy subjects. RA patients were found to have a significantly higher degree of IgE-IgG switching compared to healthy individuals (*P*=1.32e-02, **Fig. 6b**). At the clinical phenotype level, we found no evidence of heterogeneity of isotype class switching in RA phenotypes (**Table S26**).

After the initiation of the immune response, B cells increase affinity to antigens by undergoing somatic hypermutation in the V segment genes. To determine whether the level of BCR affinity is modified in RA, we analyzed the somatic hypermutation profile of IgM, IgA, IgG, IgD and IgE IGH isotypes as well as in IGL and IGK chains. The degree of somatic hypermutations in IGL (*P*=1.31e-02), IgM (*P*=1.26e-02) and IGK (*P*=2.04e-02) chains was found to be significantly higher in RA compared to controls (**Fig. 6c**). Stratifying the analysis by clinical phenotype, the IgE isotype showed a distinct somatic hypermutation profile in patients with high and low disease activity (*P*=4.36e-02, **Table S26**). RA patients also showed a significantly higher percentage of mutated IgD/M than healthy subjects (*P*=4.31e-03, **Fig. 6d**). In the clinical phenotype analysis (**Table S26**), we detected that ACPA-positive and ACPA-negative patients differ on the percentage of mutated IgD/M (*P*=4.49e-02).

### Multi-chain AIRR predicts RA

To assess if the information residing in the AIRR of RA patients could be translated to information directly applicable in medical practice, we evaluated the utility of AIRR sequencing as a tool for disease diagnosis. Using a leave-one out cross-validation strategy, we found that, rather than a single AIRR measure being useful for prediction, aggregating information from different AIRR properties lead to an optimal disease classifier (**Table S27**). Combining usage information from the seven receptor chains and each of the 298 V gene segments, we built a disease classifier with a 95.2% accuracy (**Fig. 7a-b**). Among these predictor variables, IGKV2-28, TRAV20 and TRBV25-1 segments were found to be the AIRR features that contributed most to RA classification (**Fig. 7c**).

**FIGURE 7.**
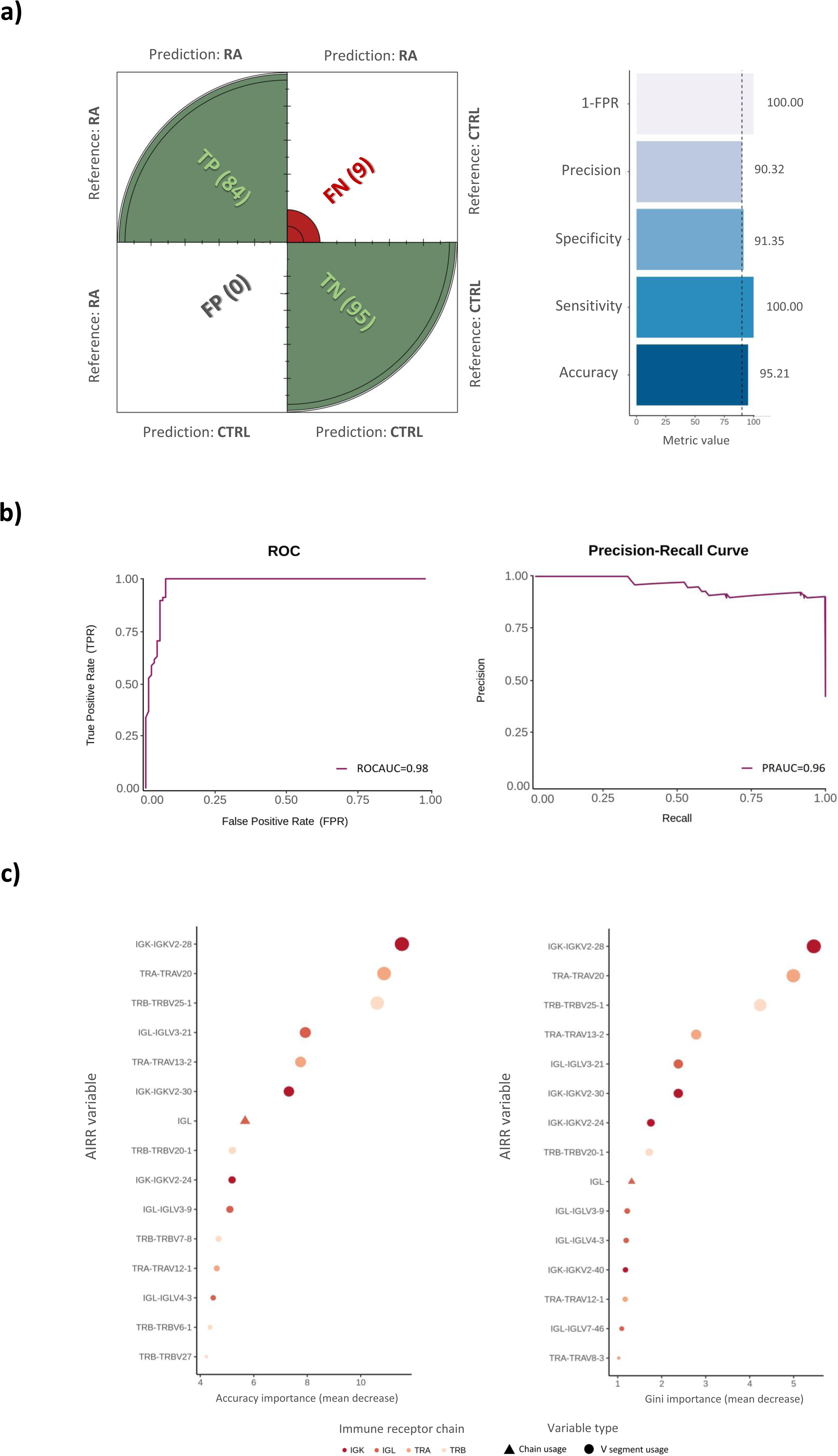
Development of a multi-chain AIRR predictor for rheumatoid arthritis. a) Predictive performance of the optimal classifier that aggregates chain and gene segment usage information. On the left side, the graphical representation of the confusion matrix is shown. The number of individuals that were predicted as patients or controls is shown between parenthesis. On the right side, the value of the five metrics used for evaluating the predictive performance is shown. Abbreviations: CTRL, controls; RA, rheumatoid arthritis; FN, false negatives; FP, false positives; FPR, false positive rate; TP, true positives; TN, true negatives. b) Receiver operating characteristic and precision-recall curves illustrating the diagnostic ability of the multi-chain AIRR classifier for rheumatoid arthritis. Abbreviations: ROC, receiver operating characteristic; ROCAUC, are under the receiver operating characteristic curve; PRAUC, are under the precision-recall curve. c) Identification of the 15 AIRR predictor variables that contributed most to the diagnosis of patients with rheumatoid arthritis in accordance with the mean decrease in accuracy and Gini importance measures.

We also interrogated the potential of AIRR-seq information for predicting clinically relevant phenotypes in RA, particularly response to therapy. We found that while the percentage of IgD/M mutated at baseline only predicted 56.25% of non-responder patients to TNFi therapy (**Table S27**), the specificity increased substantially (68.75%) when only patients with extreme clinical responses (i.e., non- and good-responders) were included in the analysis (**Table S27**). The power of AIRR-seq information for predicting other clinical features was found to be variable, ranging from a low accuracy for disease activity (57.1%) to improved accuracies for RF (69.8%) and ACPA (74.7%) positivity ( **Table S27**).

## METHODS

### Patients and samples

A total of 86 RA patients and 104 healthy subjects were included in the present study (**Table S1**). All patients were collected from the outpatient’s clinics of the rheumatology departments of 12 Spanish University Hospitals involved in the Immune-Mediated Inflammatory Disease Consortium^38^. Controls were recruited from healthy blood donors from Spanish hospitals in collaboration with the Spanish DNA Bank. Enrolled patients fulfilled the ACR/EULAR 2020 classification criteria for RA and had more than 2 years of follow-up since diagnosis^39^. All recruited patients had an erosive disease defined as erosions in more than one joint group including hands and/or feet. The clinical response to TNF inhibition therapy was measured using the European League Against Rheumatism treatment response criteria^40^. For all patients, the Disease Activity Score (DAS28) was measured at baseline and after 12 weeks of TNFi therapy. According to the DAS28 variation and the DAS28 at the endpoint, RA patients were categorized into good (n=42), moderate (n=25), and non-responders (n=17). Good and moderate responders were grouped into a single and larger responder group. A DAS28 threshold of 5.1 at baseline was used to categorize RA patients into high (i.e., DAS28>=5.1, n=48) and moderate-low (i.e., DAS28<5.1, n=38) disease activity. All RA patients included in this cohort were Caucasian European and born in Spain. The main clinical and epidemiological characteristics of this cohort of RA patients are summarized in **Table S1**.

All the procedures were followed in compliance with the principles of the Declaration of Helsinki and all patients provided written informed consent to participate in this study. The study and the consent procedure were approved by the local Institutional Review Board of each participating center.

### Whole blood RNA extraction

Peripheral whole blood from venipuncture was dispensed into PAXgene™ Blood RNA Tubes (Qiagen). Whole blood samples in PAXgene™ Blood RNA Tubes were stored at -80°C until RNA extraction. RNA was extracted from PAXgene™ Blood RNA Tubes via the PreAnalytiX PAXgene™ blood RNA kit (Qiagen) and automated using the QIAcube.

### High-throughput sequencing of the seven receptor chains of the study population

Two independent next generation sequencing libraries covering all TCR (i.e., TRA, TRB, TRD and TRG) and BCR (i.e., IGH, IGL and IGK) chains were generated from each sample using iR-RepSeq+ 7-Chain service (iRepertoire, US). This technology allows the amplification of the seven receptors chains in an unbiased manner^30^. In this procedure, the first set of primer pairs for each V-J combination is used, allowing the extension with tags that, in turn, enable a second set of primers to be utilized for global amplification of all seven chains. This unbiased amplification was conducted in a single assay using UMI during the reverse transcription step. The introduction of UMI to distinguish between single RNA molecules during cDNA synthesis was essential to minimize the impact of PCR duplicates and sequencing errors. All necessary reagents for amplification and purification are preloaded into the cassette. Extracted RNA (1,000 ng) with an appropriate volume of RT mix and nuclease-free water were added into the cassette, which was processed by the iR-Processor. The instrument can automatically set up and carry out all amplification and purification. Briefly, RT is performed using Qiagen OneStep RT-PCR mix (Qiagen). First-strand cDNA was selected, and remnant primers were removed by SPRIselect bead selection (Beckman Coulter) followed by the second round of binding and extension with the V-gene primer mix. After binding and extension, SPRIselect beads were used to purify the first and second strand synthesis products. Library amplification was performed with a pair of primers that are specific for communal sites engineered onto the 5’ end of the C- and V-primers used in first and second-strand synthesis. The final constructed library includes Illumina dual index sequencing adapters, a 10-nucleotide UMIs, and an 8-nucleotide internal barcode associated with the C-gene primer. Amplified libraries were multiplexed and pooled for sequencing on the Illumina NovaSeq platform with a 500-cycle kit (250 paired-end reads) at Hudson Alpha Institute for Biotechnology (Huntsville, AL, USA). The output of the immune receptor sequence covers within the first framework region through the beginning of the constant region, including the CDR3 hypervariable region.

### AIRR-seq data processing

Illumina paired-end reads were demultiplexed and collapsed by 8 base pairs barcode and 10 base pairs UMI tag using the default *MIG* size threshold set by MiGEC v1.2 software^41^. In order to assemble the full TCR/BCR sequences, the resulting UMI-collapsed fastq files were mapped to the V, D, J and C reference sequences from the International ImMunoGeneTics (IMGT) database using the MiXCR software (v3.0.14) with the clustering parameters turned off^42, 43^. Sequences that were out-of-frame or contained stop codons were filtered out from the analysis. The resulting AIRR-seq data contained sequences from the seven chains that cover the receptor sequence spanning from the FR1-IMGT to FR4-IMGT region, as well as the constant region of the receptor. Sequencing information contained within the constant region of IGH was used to compute the isotype-specific features described in the “*Isotype information contained within the constant region of IGH”* section. In order to increase the statistical power of the present analysis, AIRR-seq data from each sample and its corresponding replicate were merged by the CDR3 nucleotide sequence. The total number of reads and UMIs obtained at each stage of the pipeline are summarized in **Table S2**.

### Genome-wide genotyping and *HLA* imputation

All individuals included in the case-control cohort from the Spanish population (n=190) were genome-wide genotyped using Illumina Global Screening Array-24 Multi Disease version 2.0 (Illumina, USA) at Life and Brain Center (Bonn, Germany). Genotype calling and quality control analyses were performed using GenomeStudio (Illumina, USA) and PLINK softwares, respectively^44^. Two samples showing a call rate <0.95 were excluded from the study and, therefore, samples from 84 RA patients and 104 healthy subjects were finally available for analysis. Using GWAS data from our study population, we performed the imputation of the HLA classical alleles and amino acid polymorphisms. HLA imputation was conducted using the SNP2HLA V.1.0.3 software^45^.

### Definition of AIRR variables

#### Clone

In the present study, clones were defined by the CDR3 amino acid sequences. Accordingly, the number of unique clones detected within the AIRR of a particular individual corresponds to the total number of unique CDR3 amino acid sequences detected in this individual.

#### Clone publicity

In the present study, the publicity degree of each clone was defined as the number of individuals where the clone is detected.

#### Meta-clone

Groups of clones showing a high similarity at the CDR3 amino acid sequence level.

#### K-mer

K-mers are groups of amino acids of length K contained within the CDR3 amino acid sequence^46^. Here, the CDR3 amino acid sequence of each clone was broken into overlapping k-mers of 4 amino acids of length.

#### Diversity

Diversity is one of the most commonly used metrics to characterize and quantify the vastness of the AIRR. Here, diversity was calculated at the chain level according to the observed frequency of clones within an individual’s AIRR. Given the lack of a single measure that efficiently captures all diversity features and the availability of different measures that provide complementary information on the AIRR diversity^47^, we computed five common diversity measures to characterize the diversity of the AIRR in RA: Diversity 20 (D20), Diversity 50 (D50), Gini-Simpson, Inverse-Simpson and Shannon entropy. D20 and D50 measures represent the number of dominant unique clones which make up 20% and 50% of the total clones, respectively. Gini-Simpson measure is computed as the unity minus the Simpson’s index and indicates if the repertoire is oligoclonal with a few highly frequent clones (i.e., low values) or polyclonal with a similar representation of each clone (i.e., high values). For the present analyses, the Gini-Simpson index was log transformed. Inverse-Simpson index was obtained using the weighted arithmetic mean to quantify the average proportional abundance of clones, thereby indicating an even distribution of clones (i.e., high values) or clonal expansion (i.e., low values). Finally, the Shannon entropy was computed to account for both the unique clone number (i.e., repertoire richness) and the clone relative abundance (i.e., repertoire evenness).

#### Clone length

The length of each clone was determined by the number of amino acids that are present in its CDR3 amino acid sequence.

#### Chain usage

In order to determine the relative mRNA expression of each immune receptor chain, we computed the chain usage measure. For each individual, the usage of each immune receptor chain was defined as the proportion of chain UMIs.

#### V and J gene segment usage

The genomic rearrangement between V and J gene segments determines the V(D)J junction, which includes the CDR3 hypervariable region. The AIRR of an individual can be therefore described not only by the frequency of CDR3 sequences, but also by the frequencies of each V and J segments as well as the frequency of each VJ combination. Accordingly, we computed the usage of each V, J and VJ segments as their frequency within the AIRR of each individual.

#### Isotype information contained within the constant region of IGH

In order to characterize the molecular signatures that characterize IGH isotypes in RA patients, we have computed four informative measures of the antibody functional role. Using sequencing information contained within the constant region of IGH, we have computed the isotype use (i.e., percentage of unique VDJ sequences per isotype), isotype class switching recombination index (i.e., frequency of unique VDJ regions expressed as two isotypes) and both the percentage of mutated IgD/M and degree of somatic hypermutations (i.e., average number of mutations per unique VDJ region sequence) as previously described^23^. In order to quantify these isotype-specific features, the *exportAlignments* function of MiXCR software was used to get maximum length sequences. Given the small number of occurrences in which an IgE primer picked up an IgG sequence, which were incorrectly called as IgE by the MiXCR pipeline, we conducted an *ad hoc* post-processing step that corrected this spurious isotype calling by realigning the reported IgE sequences with the Smith-Waterman algorithm^48^.

### Statistical analysis

#### Influence of technical factors and epidemiological variables on AIRR properties

We first investigated the impact that both technical factors (i.e., sequencing depth, sequencing plate, RNA integrity and RNA concentration) and epidemiological variables (i.e., age and gender) have on the main AIRR properties (**Fig. S1**). After detecting technical factors showing a high degree of collinearity, we tested the influence of non-redundant technical variables and epidemiological variables on each AIRR measure. To ensure that our analyses are not biased by technical aspects, technical factors influencing AIRR measures (r^2^>0.25, *P*<0.05) were considered when the epidemiological variables were tested for association with these measures. The association between quantitative variables was assessed using the Pearson correlation. The association between categorical variables with two and more than two labels was determined using the Wilcoxon and Kruskal-Wallis tests, respectively. The complete list of associations is detailed in **Fig. S1**. The downstream AIRR-seq analyses presented in the present study were adjusted by age and gender as well as in accordance with the statistical association observed between each technical variable and AIRR measure.

#### Clone publicity analysis

Since the degree of clone sharing between individuals is strongly influenced by the number of individuals included in each group of interest, we performed a rarefaction analysis to compare the clone publicity between groups with an unbalanced sample size.

#### V and J gene segment usage analysis

In order to characterize the gene segment usage profile of our case-control cohort, gene segments showing a usage <5% were excluded from the study. Subsequently, we conducted a principal component analysis to determine the potential of gene segment usage to differentiate RA patients from healthy individuals. The two components explaining the largest fraction of the gene segment usage variance were selected for the analysis.

#### Clone length comparative analysis

We conducted a multi-step statistical analysis to detect: i) shifts in the clone length distribution between two conditions, and ii) the amino acid positions at which the differential pattern starts. To identify shifts in the clone length distribution between the groups of interest, the frequency of clone abundance was compared using the Wilcoxon test. Differences in the abundance of clones with a particular length of the CDR3 amino acid sequence between two conditions were evaluated using Fisher’s exact test. To better characterize the amino acid positions at which the differential pattern starts, we computed both the empirical cumulative density distribution function and the quantile-quantile plots. These graphical representations were used to determine if the clone length frequencies observed in two different conditions follows the same probability distribution.

#### Baseline association analysis

In order to identify AIRR-seq variables associated with a trait of interest, we used logistic and negative binomial regression models according to the distribution followed by the AIRR-seq variable analyzed (**Table S28**). Epidemiological variables and technical factors like sequencing depth were included as covariates in all statistical models.

Given the vastness of the AIRR and the complexity of translating repertoire information contained in a few milliliters of blood to the whole individual’s AIRR (i.e., sampling bias), discovering clones associated with complex traits is highly challenging. The little clone sharing (i.e., clone publicity) detected between our study samples leads to an extremely zero-inflated clone distribution (**Fig. S8, Fig. S9**). This particularity of the AIRR-seq data represents a major analytical challenge to identify disease-associated clones with high confidence^24, 49^. To identify clones that are over- or under-represented in disease conditions, in the present study we adapted the Hurdle model^50^. This model is commonly used in single cell RNA-seq studies and allows the comparison of extremely zero-inflated clone abundance distributions^51^. It is a two-part generalized linear model that simultaneously models the rate of clone abundance over the background of various clones and the positive clone abundance mean. This modelling allows the inclusion of covariates in both the discrete and continuous parts of the model allowing the adjustment of the model for confounding factors. To adjust our model for important factors of AIRR-seq analyses, we designed and implemented the Clone Detection Rate (CDR) measure that acts as a proxy for both technical (e.g., sampling bias) and biological factors (e.g., RNA volume and extrinsic factors other than the clinical phenotype of interest) that globally influence clone abundances. The CDR was computed as the standardized proportion of clones that are detected in each sample. Together with the statistical properties of the Hurdle model, the adjustment for the variability of AIRR-seq data captured by the CDR makes the Hurdle model more effective at controlling the impact of the technical factors and reducing the false positive rate than other analytical strategies commonly used in AIRR-seq analyses (e.g., Fisher’s exact test). Before using the Hurdle model, clone counts were adjusted for the library size, multiplied per million and converted to the logarithmic scale. After excluding those clones detected in <1% of samples (**Table S29**), the association of each clone with disease conditions was tested using the Hurdle model adjusted for CDR, age and gender. The False Discovery Rate (FDR) method was used to account for multiple testing^52^.

The stringent multiple testing correction that must be applied to the association analysis at the single clone level can reduce the power to identify disease-relevant clones. This statistical power limitation raises the need of developing new approaches to identify disease-associated clones. Given that clones with high sequence similarity are likely to recognize similar antigens^34^, we exploited the similarity information across CDR3 amino acid sequences using network analysis to identify meta-clones associated with RA (**Fig. 4e**). Our meta-clone approach starts with the construction of the CDR3 sequence similarity network of millions of clones that follow an extremely zero-inflated frequency distribution. This is built using the *mutation.network* function of the tcR R package^53^. In this network, edges connect those clones that differ by at most one amino acid. Communities of highly connected clones (i.e., meta-clones or groups of clones with a high CDR3 amino acid sequence similarity) are subsequently identified by directly optimizing a modularity score with the *cluster.fast.greedy* function of the igraph R package^54^. The expression of each meta-clone is then computed by aggregating the expression values of the community of highly similar clones. Finally, the association of each meta-clone (i.e., detected in >1% of samples) with disease conditions is tested using the Hurdle model adjusted for age, gender and detection rate as previously described (**Table S29**). The same analytical approach was used to test the association between k-mers and disease conditions (**Table S29**).

#### Longitudinal association analysis

In order to evaluate the impact that TNFi drugs have on the main AIRR-seq features, we implemented a logistic and negative binomial regression (i.e., determined by the distribution of the AIRR-seq variable analyzed, **Table S28**) using a mixed effect model. Like in the baseline association analysis, sequencing depth and epidemiological variables were included as covariates in the mixed effect models.

Similar to the baseline association analysis, studying the dynamics of the AIRR at the resolution of single clones must allow to distinguish true differences in clone frequencies from experimental noise. To address this analytical challenge in the study of the impact of TNFi therapy at the single clone level, we used NoisET^31^. Briefly, NoisET is a newly developed Bayesian approach that exploits the information of each sample and replicate for learning a noise model that accounts for read counts variability due to sampling, library preparation and expression noises. Following the method’s guidelines, we selected the negative binomial model for learning the experimental and technical noise. According to the AIRR-seq data required for the probabilistic method implemented in NoisET, we used the CDR3 nucleotide sequence of each clone to infer the noise model that allowed the unbiased identification of contracted and expanded clones after TNFi therapy.

Differences in the number of significantly contracted and expanded clones between BCR and TCR chains were determined using Fisher’s exact test. The effect of TNF inhibition on contracted and expanded clones was compared using the Wilcoxon test. The same statistical test was used to compare the effect of TNF inhibition on contracted and expanded clones between BCR and TCR chains.

### Grouping lymphocyte interaction by paratope hotspots

To identify groups of TRB clones with shared specificity that are expanded in RA, we used the grouping of lymphocyte interactions by paratope hotspots (GLIPH) algorithm version 2^36^. GLIPH2 method is designed to cluster clones from the TRB chain according to their probability of binding the same Major histocompatibility complex-restricted peptide antigen. In the three-step algorithm implemented in GLIPH2, global similarity and local similarity are first computed. While the former is based on differences on the CDR3 sequence of up to one amino acid, the latter is based on the sharing of amino acid motifs among CDR3 sequences (i.e., >10-fold enriched and <0.001 probability of occurring at this level of enrichment in the naive repertoires used as reference). The global and local similarities are then used to construct clusters of TRB clones with shared specificity. The enrichment of each cluster of TRB clones in CDR3 length, clonal expansions, shared *HLA* alleles in subjects, motif significance and cluster size is next tested. Finally, the resulting probabilities of each enrichment are combined and summarized in a single score. Given that TRB clones quantified in our study population were determined from bulk RNA and no information from T cell types was available, the enrichment of the TRB repertoire was tested against a naive repertoire of both CD4+ and CD8+ T cells. To avoid false positive results, TRB clusters composed of less than three clones and detected in less than three individuals were removed from the analysis.

### Genetic association between HLA alleles and RA risk

The imputed HLA alleles were tested for association using a logistic regression model. In order to increase the power of identifying HLA alleles associated with RA risk in the Spanish population, we analyzed a case-control cohort of 1,191 RA patients and 1,558 healthy controls that we had previously collected^55^. The genetic association results were adjusted for multiple testing using the FDR method^52^.

### Estimating the AIRR from bulk RNA-Seq data

In order to obtain the immune repertoire from the seven receptor chains that are expressed in blood and synovial tissue of RA patients^32^, we leveraged the information provided by TCR and BCR transcripts that are present in a publicly available bulk RNA-seq dataset using MiXCR software^43^. MiXCR aligns paired-end reads to a built-in library of V, D, J and C gene sequences and, subsequently, information from the alignment of the two reads is aggregated to increase the gene assignment accuracy. After correcting for PCR and sequencing errors, identical and homologous reads are assembled into clones. In order to maximize the quantitative information available for analysis, low-quality reads are rescued by mapping them to previously assembled high-quality clones.

### Clone characterization

#### Amino acid sequence similarity

In order to quantify the similarity between amino acid sequences, we implemented two analytical approaches. The sequence similarity among a given set of clones was determined using a network analysis strategy. This analytical strategy consisted on building a CDR3 amino acid sequence similarity network, where each clone is represented as a node and edges connect CDR3 amino acid sequences differing by at most one amino acid. The degree centrality value of the sequence similarity network was used to estimate the similarity among the set of clones. The similarity among a given set of smaller amino acid sequences (i.e., k-mers) was determined using the Levenshtein distance metric, which represents the minimum number of characters that differs between two strings of characters^56^. Unlike the commonly used Hamming distance, which is limited to count character substitutions between two sequences of the same length^57^, the Levenshtein distance metric considers also the presence of character insertions/deletions and, therefore, it can provide additional insights into the characterization of the immune repertoire^58^.

#### Biochemical profile of the CDR3 amino acid sequence

The sequence profile of a given set of CDR3 amino acid sequences was determined using the ClustalW alignment algorithm with the R package msa^59^. The sequence logo of the resulting alignment was built using the probability method provided by the R package ggseqlogo^60^. Each amino acid was colored according to its most characteristic biochemical feature (i.e., acidic, basic, hydrophobic, neutral or polar). In order to refine the biochemical characterization of a given set of CDR3 amino acid sequences, we studied additional biochemical properties using the *Peptides* R package. To determine whether the biochemical profile of expanded and contracted clones differs between responder and non-responder patients to TNFi therapy, the enrichment in each biochemical property was compared using a logistic regression model adjusted for the length of the CDR3 amino acid sequence. After weighting the biochemical profile of the significantly expanded or contracted clones by the magnitude of the frequency change, the same analytical approach was used to quantify the global effect that TNF inhibition exerts on the AIRR biochemical profile that characterizes responder and non-responder patients.

#### Comparison against a random repertoire

In order to determine whether a given set of CDR3 amino acid sequences are characterized by a particular biochemical property or a higher sequence similarity than expected by chance, we have used a resampling strategy. Using this analytical approach, the statistical significance of the feature (e.g., sequence similarity) was computed by contrasting the observed metric in a given set of clones against the null distribution obtained after sampling multiple times (i.e., here >1,000 and 1,000 for sequence similarity and biochemical properties, respectively) a random set of clones from the same size and receptor chain. After obtaining the statistical significance of enrichment and depletion in a particular biochemical feature, both statistics were log-transformed (i.e., -log10 and log10 for enrichment and depletion p-values, respectively) and subsequently aggregated into a single biochemical enrichment score (BES).

### Development of the multi-chain AIRR predictor

To investigate whether AIRR molecular data could be translated into information of clinical utility, we built and tested predictor models. Multi-chain AIRR features (n=4,521 variables) and HLA genetic information (n= 6,407 genetic variants) from 84 RA patients and 104 control individuals were evaluated as predictors (**Table S30**). The predictive power of AIRR information was separately analyzed for each AIRR feature as well as aggregating information from different AIRR features. The analytical approach selected for the present predictive analysis was composed by the random forest algorithm and leave-one-out cross validation strategy. To avoid biased prediction resulting from the analysis of imbalanced classes (e.g., more TNFi responders than non-responders), we used the random forest algorithm with down-sampling (i.e., sampling the majority class to make their frequencies closer to the minority class). The predictive performance was determined using five discriminative measures: i) accuracy, ii) sensitivity, iii) specificity, iv) precision and v) false positive rate. For the most powerful predictor both the area under the receiver operating characteristic and the area under the precision-recall curve were computed.

## DISCUSSION

The present work represents the first comprehensive study of the seven immune receptor chains in autoimmunity. Using a novel high-throughput sequencing method and three complementary study designs, our analytical pipeline reveals a profound heterogeneity in the AIRR of RA. We demonstrate that AIRR features like diversity are not equally distributed between the seven immune receptor chains and disease conditions. Many of the AIRR associations with disease and clinically relevant phenotypes that we have found have not been previously described, including those related with the longitudinal evolution of the AIRR under TNFi treatment. This study provides a deep quantitative analysis on the variability of the adaptive immune response in RA, and it is a step forward to develop new precision medicine strategies based on circulating B and T cell receptor repertoires.

The quantitative analysis performed here has allowed to characterize, for the first time, the main AIRR features of RA at the resolution of the seven receptor chains. We have demonstrated that the low AIRR diversity detected in RA patients is restricted to the B cell compartment. Our findings confirm the expansion of BCR clones in RA, which is consistent with the previous identification of dominant B cell clones both in circulation and in the RA synovium^61, 62^. We also demonstrate that therapeutic inhibition of TNF is able to restore the BCR chain diversity, raising it to the levels observed in healthy individuals. A previous longitudinal study on anti-CD20 therapy (rituximab) in RA showed the impact that biological therapies have on BCR diversity^29^. However, this is an expected finding in a drug that directly depletes B cells. Instead, the strong impact of TNFi therapy on IGH, IGK and IGL diversity reported here supports a novel downstream effect of the drug’s mechanism of action.

The stratification of the longitudinal analysis by treatment efficacy contributed to better understand the differences in AIRR diversity between TNFi responders and non-responders. We found that response to treatment is associated with a bigger increase in diversity, and that this is mainly driven by the reduction of BCR clones that share a specific biochemical profile. The latter finding suggests that TNFi responders are reacting against a different group of autoantigens than non-responders and, therefore, antibody-mediated immune mechanisms could also be different between the two groups. The recent identification of subtypes of macrophages shaping synovial membrane heterogeneity in RA support the presence of distinct effector responses to antigen-bound antibodies^63^. In this context, different (self) antigen-antibody complexes could elicit diverse proinflammatory mechanisms through the Fc-receptors on macrophage subtypes. Differences in sensitivity to TNFi in these activated macrophages could explain the observed heterogeneous response to this therapy in RA^64–66^.

In this work, we have characterized the TCR and BCR repertoire underlying RA by interrogating a wide variety of AIRR properties. Exploiting the multi-chain data provided by a new technology, we have detected a low abundance of IGL in the blood of RA patients. We have demonstrated that the usage of this BCR chain is higher in synovium than in blood, suggesting a strong migration of IGL-harboring B cells to the target tissue of RA. A similar finding has been recently described in COVID-19, where patients with more severe disease display an increased migration of T cells from blood to inflamed tissues like the lungs^67, 68^. Also, we have found that the multi-chain gene segment usage profile is highly distinctive between RA and control subjects. This result corroborates previous findings derived from single-TCR chain analyses, and reveals the concerted association at the chain-wide scale^28^.

Association of the immune repertoire to clinical phenotypes was variable. Some phenotypes like the presence of autoantibodies showed significant association to multiple AIRR features, while disease activity showed a more moderate impact. One of the AIRR features that was found to be most strongly associated to multiple RA phenotypes was clone length. Previous studies have found large CDR3 amino acid sequences in RA synovial membrane^61^, supporting the hypothesis that longer amino acid sequences are more likely to react against self-antigens^37^. Our results provide clear evidence that this feature is characteristic of the circulating AIRR of RA patients, and also extend the relevance of clone length to other clinical phenotypes. Indirectly, the association of large CDR3 sequences in TRD clones provides additional support to the pathogenic role for this rare T cell subset in the pathology of RA^69, 70^.

Immunoglobulin isotype class switching and somatic hypermutation delineate the functional properties of antibodies. By performing an in-depth characterization of the main isotype features, we have found a high level of IgE-IgG switching in RA. This finding is in line with recent evidence in systemic lupus erythematosus and Crohn’s disease showing also an association to this specific isotype class switching^23^. IgG but not IgE isotypes play a key role in the activation of the complement pathway and, therefore, our findings also support an increased activation of the complement system in RA^21, 71^. The lower IgD and higher IgG levels that we detected in RA compared to controls are in accordance with previous studies measuring the serological levels of these two isotypes^72^. The accumulation of somatic hypermutations detected in RA patients is also in line with recent evidence supporting that this affinity maturation step contributes to increased polyreactivity to different self-epitopes by altering key amino acid residues within the antibody paratope^73, 74^. The highly mutated IgD/M isotypes detected in ACPA-positive RA could therefore reflect the raised production of B clones that recognize citrullinated peptides in this group of patients^75^.

One of the main challenges in AIRR-seq analyses is the robust identification of specific clones associated with traits^24, 49^. The extraordinary size and diversity of the TCR and BCR repertoires severely limits the statistical power to identify trait-specific clones. To overcome this, we have adopted two new methodological strategies. First, we have introduced the use of an analytical methodology commonly used in single cell RNA-seq studies that is robust to the overabundance of zero values^50^. Second, to identify the temporal changes in clonal frequencies after three months of TNFi therapy, we have used a new Bayesian method that accounts for the clonal frequency variability that is due to sampling, library preparation and expression noises^31^. This latter approach has proven successful to identify contracted T cell clones after COVID-19 infection^27^. In the case-control analysis, we were able to confidently identify 20 IGL and 40 IGK clones associated with disease. The striking low levels of all significant clones in RA patients raise the possibility of B cell subtypes that are protective of autoimmunity. This is in line with the growing evidence on the immunosuppressive role that B regulatory cells have on autoimmune diseases^76^. Previous studies have shown the existence of PD-L1+ B regulatory cells with CD8+ T cell suppressive capacity that are reduced in RA compared to healthy controls^77^. Our results point towards a depletion of specific IGL- and IGK- harboring B regulatory cells that are needed for maintaining the immune tolerance.

The search for clonal features associated with RA at the chain-wide scale was also extended to groups of clones with similar amino acid motifs. For this objective, we developed meta-clone and k-mer analytical strategies that yielded 4 meta-clones and 100 k-mers associated with RA. The detection of k-mers spanning the amino acid sequences of clones from TCR chains that are over-represented in RA is consistent with the previous identification of highly expanded T cell clones in RA blood^78, 79^. Since the T cell repertoire is markedly shaped by variation at the *HLA* loci^80^, we also conducted TCR association analysis with HLA variants in our study cohort. Integrating this data in our analysis, we found that the cluster of CD4+ TRB clones that harbors the QDFA amino acid motif is both enriched in the RA repertoire and associated with *HLA-DRB1*15,* the class II *HLA* variant that is most strongly associated with RA risk in the Spanish population. *HLA-DRB1*15* has also been previously associated with an increased production of ACPA levels in RA^81^. Accordingly, the present findings support that part of the genetic risk to RA could be mediated by the *HLA* restriction of the hypervariable region of TRB clones that recognize citrullinated peptides. The TRB clones associated with disease risk *HLA* variants identified in our work represent a useful reference that could help to identify additional autoantigens in RA.

In the present study, we also demonstrate that AIRR-seq information residing in the seven receptor chains has a high promising diagnostic potential in RA. This finding represents an important step to overcome one of the currently unmet needs in RA, which is the lack of peripheral blood biomarkers with high accuracy for disease diagnosis^82^. While anti-cyclic citrullinated peptide antibodies have a high specificity, they lack sensitivity due to the considerable fraction of autoantibody-negative patients in RA (25-30%). The development of an accurate biomarker for early disease diagnosis in both ACPA-positive and ACPA-negative patients is therefore of major clinical interest. In particular, validation of the AIRR-seq predictor in early RA patients could substantiate earlier and thereby more efficacious therapeutic interventions.

One of the current challenges in AIRR-seq studies is to provide replication on independent populations, as it has become standard in other genomic analysis fields like genome-wide association studies^83^. With the increasing sharing and standardization of AIRR-seq data like the initiative promoted by the AIRR Community^13^, the availability of chain-wide immunosequencing data from different case-control cohorts will enable the validation of our findings before moving to the clinical setting. In the future, immune receptor profiling at the single-cell resolution will be key to refine the present findings down to the T and B cell subtype level. To date, however, the high costs that would involve sequencing the repertoire of millions of cells in large cohorts of individuals, together with the huge diversity of circulating T and B lymphocytes, makes bulk repertoire analysis a cost-efficient and highly informative approach for studying the AIRR in autoimmunity.

## DATA AVAILABILITY

The immunosequencing data from the seven receptor chains used for this study will be available for download at the website of the Rheumatology Research Group (www.urr.cat/AIRR-RA) upon acceptance. All the research findings will be also available for visualizing and browsing on the website of the research group upon acceptance. Custom R code used for analyzing the AIRR-seq data from the seven receptor chains will be publicly available at www.github.com/Rheumatology-Research-Group/AIRR-RA upon acceptance.

## Data Availability

The immunosequencing data from the seven receptor chains used for this study will be
available for download at the website of the Rheumatology Research Group
(www.urr.cat/AIRR-RA) upon acceptance. All the research findings will be also available for
visualizing and browsing on the website of the research group upon acceptance. Custom R
code used for analyzing the AIRR-seq data from the seven receptor chains will be publicly
available at www.github.com/Rheumatology-Research-Group/AIRR-RA upon acceptance.

https://www.github.com/Rheumatology-Research-Group/AIRR-RA

## ACKNOWLEDGMENTS

We thank the patients and clinical specialists collaborating in the IMID Consortium for participation. We thank David Ellinghaus and Frauke Degenhardt (Institute of Clinical Molecular Biology, Kiel University) for conducting the genome-wide genotyping of the study population. This study was funded by Instituto de Salud Carlos III through the project PI17/00019 (Co-funded by European Regional Fund “A way to make Europe”) and by the Spanish Ministry of Economy and Competitiveness (grant number: IPT010000–2010–36). The study sponsors had no role in the collection, analysis or interpretation of the data.

## AUTHOR CONTRIBUTIONS

AA, RMM, SM, JH and AJ conducted the study design and data interpretation. MLL, FB, AJM, MLGV, AE, CPG, SASF, RS, AFN, MAL, JT, AMO, JM, NP and SM participated in patient recruitment and sample acquisition. IR, RMM, WP and MB-S were involved in AIRR immunosequencing and review of the data. AA, AJ, DW and DS performed AIRR-seq data analyses. AA, SM and AJ wrote the manuscript. All authors revised the manuscript and gave final approval for its submission.

## SUPPLEMENTARY FIGURES

**FIGURE S1. Impact of technical and epidemiological variables on AIRR features.** Evaluation of collinearity among technical variables (i.e., RNA concentration, RNA integrity, sequencing depth and plate), association of technical variables with AIRR features as well as association of epidemiological variables with AIRR features (i.e., adjusted by the influencing technical factors).

**FIGURE S2. Chain-wide characterization of the AIRR diversity in rheumatoid arthritis phenotypes.** The statistical significance and effect size of the association between each diversity measure and clinical phenotype are shown. The heatmap is colored according to the effect size of the association at the chain level. Details on the direction of the effect size are provided in **Table S4**. Abbreviations: P<0.05; **, P<5.00e-03; ***, P<5.00e-04; ****, P<5.00e-05.

**FIGURE S3. Chain usage association with clinical phenotypes in rheumatoid arthritis.** The percentage of patient UMIs mapping to each immune receptor chain is separately represented by clinical phenotype.

**FIGURE S4. Graphical representation of the significant associations between IGL/IGK clones and rheumatoid arthritis.** Significant associations detected by the Hurdle or continuous models are represented using violin plots, where the clone expression is plotted separately for each phenotype. Significant associations detected by the discrete model are represented using bar plots, where the number of individuals having the clone are plotted separately for each phenotype. For all significant associations (FDR<0.05), the clone expression is also plotted against the standardized clone detection rate. Abbreviations: CDR, clone detection rate; CPM, count per million on the logarithmic scale; Cont, continuous model; CTRL, healthy individuals; Disc, discrete model; P, p-value; RA, rheumatoid arthritis.

**FIGURE S5. Graphical representation of the significant associations between TRA/TRB/IGL/IGK k-mers and rheumatoid arthritis.** Significant associations detected by the Hurdle or continuous models are represented using violin plots, where the k-mer expression is plotted separately for each phenotype. Significant associations detected by the discrete model are represented using bar plots, where the number of individuals with clones harboring the k-mer are plotted separately for each phenotype. For all significant associations (FDR<0.05), the k-mer expression is also plotted against the standardized k-mer detection rate. Abbreviations: Cont, continuous model; CPM, count per million on the logarithmic scale; CTRL, healthy individuals; Disc, discrete model; KDR, k-mer detection rate; P, p-value; RA, rheumatoid arthritis.

**FIGURE S6. Pairwise dissimilarity matrix among the amino acid sequences of TRA/TRB/IGL/IGK k-mers associated with rheumatoid arthritis.** K-mers over- and under-represented in rheumatoid arthritis are represented in dark and light blue, respectively. The dissimilarity index was computed using the Levenshtein distance measure so that the higher the Levenshtein distance, the higher the dissimilarity index between two k-mer sequences. Abbreviations: Cont, continuous model; Disc, discrete model; RA, rheumatoid arthritis.

**FIGURE S7. Graphical representation of the association between the length of the CDR3 amino acid sequences and rheumatoid arthritis.** The results of the case-control, case-case (i.e., association analysis with clinical phenotypes in rheumatoid arthritis) and longitudinal analysis (i.e., baseline vs. week 12 and baseline vs. week 12 stratified by clinical response) are provided at the chain level. Shifts in the clone length distribution and the statistical significance of the difference in the abundance of clones with a particular length of the CDR3 amino acid sequence between the two indicated conditions are shown on the left side. This plot also shows the summary statistics detected for each condition. In the middle, the empirical cumulative density distribution of the abundance of CDR3 amino acid sequences is shown separately for each condition. On the right side, the empirical quantile-quantile plot computed for the two indicated conditions is shown. Abbreviations: P, p-value of the Wilcoxon test; *, p-value<0.05 in the Fisher test; **, p-value<0.005 in the Fisher test.

**FIGURE S8. Clonality profile of the samples included in the present study.** Density distribution resulting from the analysis of the clone frequency at the sample and chain levels.

**FIGURE S9. Clone publicity profile of the study population.** Graphical representation of the percentage of clones that are shared by more than two individuals (i.e., publicity degree) at the chain level. The clone publicity profile is separately shown for healthy individuals and rheumatoid arthritis patients. For each condition, the highest number of individuals sharing a clone is annotated on the right side. Abbreviations: CTRL, healthy individuals; N, sample size; RA, rheumatoid arthritis.

## SUPPLEMENTARY TABLES

**TABLE S1. Main clinical and epidemiological characteristics of the study population.** All RA patients, RA patients at baseline, RA patients after 12 weeks of TNFi therapy and control individuals are separately characterized. Abbreviations: N, sample size; NA, not applicable; M, mean; RA, rheumatoid arthritis.

**TABLE S2. Summary of the quantitative AIRR-seq information available for analysis.** The number of UMIs, clones, UMIs per sample and clones per sample are provided separately for each of the seven immune receptor chains. The total number of successfully assembled and demultiplexed reads is provided at the sample level. Abbreviations: N, number; M, mean; RA, rheumatoid arthritis.

**TABLE S3. Diversity association with rheumatoid arthritis.** For each immune receptor chain and diversity measure, both the statistical significance and effect size of the association are shown.

**TABLE S4. Diversity association with clinical phenotypes in rheumatoid arthritis.** The statistical significance and effect size of the association between each diversity measure and clinical phenotype are shown. The information is presented at the chain level.

**TABLE S5. Diversity association with TNFi therapy in rheumatoid arthritis.** For each immune receptor chain and diversity measure, both the statistical significance and effect size of the longitudinal association is shown.

**TABLE S6. Diversity variation association with TNFi therapy in rheumatoid arthritis stratified by clinical response.** For each immune receptor chain and diversity measure, both the statistical significance and effect size of the longitudinal association is separately shown for responders and non-responders to TNFi therapy.

**TABLE S7. Summary statistics of the single clone analysis at the longitudinal level.** For each chain, the number of clones that are significantly contracted and expanded after 12 weeks of TNFi therapy is shown. Detailed information on the CDR3 amino acid sequence of the significant clones is provided at the sample level. Abbreviations: N, number of clones; NA, not applicable; P, P-value.

**TABLE S8. Summary statistics of the CDR3 amino acid sequence similarity analysis among all contracted and expanded clones.** For each chain, the number of permutations used to determine the statistical significance of the sequence similarity is shown together with the observed and permuted degree centrality values. Abbreviations: N, number of permutations; M, mean.

**TABLE S9. Summary statistics of the CDR3 amino acid sequence similarity analysis among contracted and expanded clones detected in responder and non-responders to TNF inhibition.** For each chain, the number of permutations used to determine the statistical significance of the sequence similarity is shown together with the observed and permuted degree centrality values. Abbreviations: N, number of permutations; M, mean.

**TABLE S10. Chain usage association with TNFi therapy in rheumatoid arthritis.** The statistical significance and effect size of the longitudinal association is shown for each of the seven immune receptor chains.

**TABLE S11. Gene segment usage association with rheumatoid arthritis.** The summary statistics resulting from the association between each V, J and VJ gene segments with rheumatoid arthritis are shown. Abbreviations: FDR, false discovery rate.

**TABLE S12. Gene segment usage association with clinical phenotypes in rheumatoid arthritis.** The summary statistics resulting from the association between each V, J and VJ gene segments with TNFi response, disease activity, ACPA and RF phenotypes are shown. Abbreviations: ACPA, anti-citrullinated protein antibodies; FDR, false discovery rate; RF, rheumatoid factor.

**TABLE S13. Gene segment usage association with TNFi therapy in rheumatoid arthritis.** The summary statistics resulting from the longitudinal association between each V, J and VJ gene segments with TNFi therapy are shown. Abbreviations: FDR, false discovery rate.

**TABLE S14. Summary statistics of the single clone association analysis with rheumatoid arthritis.** The statistical significance of the association between each single clone and rheumatoid arthritis is shown at the chain level. For each clone, the statistical significance determined by the Hurdle, continuous and discrete models is provided. Abbreviations: FDR, false discovery rate.

**TABLE S15. Summary statistics of the CDR3 amino acid sequence similarity analysis on the single clones associated with rheumatoid arthritis.** The number of permutations used to determine the statistical significance of the sequence similarity among the disease-associated IGK and IGL clones is shown together with the observed and averaged degree centrality values. Abbreviations: N, number of permutations; M, mean.

**TABLE S16. Summary statistics of the single clone association analysis with clinical phenotypes in rheumatoid arthritis.** The statistical significance of the association between each single clone and response to TNFi therapy, disease activity, RF and ACPA phenotypes is shown at the chain level. For each clone, the statistical significance determined by the Hurdle, continuous and discrete models is provided. Abbreviations: FDR, false discovery rate.

**TABLE S17. Summary statistics of the candidate analysis between disease-associated IGK/IGL clones and clinical phenotypes in rheumatoid arthritis.** The statistical significance of the association of each single clone is shown separately for response to TNFi therapy, disease activity, RF and ACPA phenotypes. Abbreviations: FDR, false discovery rate.

**TABLE S18. Summary statistics of the meta-clone association analysis with rheumatoid arthritis.** The statistical significance of the association between each meta-clone and rheumatoid arthritis is shown at the chain level. For each meta-clone, the statistical significance determined by the Hurdle, continuous and discrete models is provided. Abbreviations: FDR, false discovery rate.

**TABLE S19. Summary statistics of the meta-clone association analysis with clinical phenotypes in rheumatoid arthritis.** The statistical significance of the association between each meta-clone and response to TNFi therapy, disease activity, RF and ACPA phenotypes is shown at the chain level. For each meta-clone, the statistical significance determined by the Hurdle, continuous and discrete models is provided. Abbreviations: FDR, false discovery rate.

**TABLE S20. Summary statistics of the k-mer association analysis with rheumatoid arthritis.** The statistical significance of the association between each k-mer and rheumatoid arthritis is shown at the chain level. For each k-mer, the statistical significance determined by the Hurdle, continuous and discrete models is provided. Abbreviations: FDR, false discovery rate.

**TABLE S21. Summary statistics of the amino acid sequence similarity analysis on the k-mers associated with rheumatoid arthritis.** Levenshtein distance observed among the TRA, TRB, IGL and IGK k-mers associated with rheumatoid arthritis. The number of permutations used to determine the statistical significance of the sequence similarity is shown together with the observed and averaged Levenshtein distance values. Abbreviations: N, number of permutations; M, mean.

**TABLE S22. Summary statistics of the candidate analysis between disease-associated TRA/TRB/IGK/IGL k-mers and clinical phenotypes in rheumatoid arthritis.** The statistical significance of the association of each k-mer is shown separately for response to TNFi therapy, disease activity, RF and ACPA phenotypes. Abbreviations: FDR, false discovery rate.

**TABLE S23. Summary statistics of the k-mer association analysis with clinical phenotypes in rheumatoid arthritis.** The statistical significance of the association between each k-mer and response to TNFi therapy, disease activity, RF and ACPA phenotypes is shown at the chain level. For each k-mer, the statistical significance determined by the Hurdle, continuous and discrete models is provided. Abbreviations: FDR, false discovery rate.

**TABLE S24. Clusters of TRB clones that are enriched in the repertoire of patients with rheumatoid arthritis and healthy individuals compared to a naive repertoire.** Summary statistics of the TRB clusters’ enrichment in the immune repertoire of rheumatoid arthritis patients and healthy controls compared to a naive repertoire of CD4+ and CD8+ T cells (P<0.1). The name of the most representative amino acid motif of each TRB cluster is based on the nomenclature used by the GLIPH2 algorithm. Detailed information on the clones that belong to each TRB cluster is also provided, including the number of unique clones, CDR3 amino acid sequences as well as V and J gene segments. Abbreviations: N, sample size, FDR, false discovery rate.

**TABLE S25. Genetic association between HLA alleles and rheumatoid arthritis risk in the Spanish population.** The frequency of each HLA allele in both the case and control populations is shown. The statistical significance and effect size of each association are also provided. Abbreviations: CTRL, control individuals; HLA, human leukocyte antigen; OR, odds ratio; P, p.-value; RA, rheumatoid arthritis.

**TABLE S26. Association of isotype-specific features with clinical phenotypes in rheumatoid arthritis.** The summary statistics (i.e., significance and effect size) of the isotype percentage, isotype class switching, somatic hypermutations and percentage of mutated IgD/M associations with disease clinical phenotype are shown. Abbreviations: P, p.-value; RA, rheumatoid arthritis.

**TABLE S27. Predictive performance of the multi-chain AIRR predictors evaluated in the present study.** The predictive performance of each multi-chain AIRR predictor is determined using the accuracy, sensitivity, specificity, precision and false positive rate measures. For each multi-chain AIRR predictor, the number of individuals that were properly classified is also shown. The AIRR feature column indicates if the multi-chain AIRR predictor was built using information from a single AIRR-feature or aggregating information from different AIRR features. Information from different AIRR features was aggregated according to different thresholds of accuracy when the sample size of the classes tended to be balanced (i.e., predictor of rheumatoid arthritis diagnosis and disease activity). The aggregation of information from different AIRR features was based on different specificity thresholds when the clinical interest was to identify the minor class (i.e., predictors of TNFi response, ACPA and RF phenotypes). The predictive performance of redundant multi-chain AIRR predictors obtained after applying different accuracy or specificity thresholds is not shown.

**TABLE S28. Detailed information on the statistical test used to determine the association between each AIRR feature and clinical phenotype of interest.** This information is provided for both the baseline and longitudinal analyses.

**TABLE S29. Main characteristics of the single-clone, meta-clone and k-mer association analyses with rheumatoid arthritis and clinical phenotypes.** For each analysis, the number of UMIs, clones, meta-clones, k-mers as well as the clone, meta-clone and k-mer detection rate per sample are shown at the chain level. The number of clones, meta-clones and k-mers detected in >1% of the samples that were included in the association analyses is also provided. Abbreviations: M, mean; N, sample size.

**TABLE S30. Complete list of the AIRR variables used for developing multi-chain AIRR predictors.** Detailed information on the 4,521 AIRR variables and 6,407 HLA genetic variants from the present study population that were available for analysis.

