## Supplementary Information for "Seven chain adaptive immune receptor repertoire analysis in rheumatoid arthritis: association to disease and clinically relevant phenotypes"

**Supplementary figures can be accessed through the following links:**

http://urr.cat/Supplementary_Information_AIRR_RA/Figure_S1.pdf

http://urr.cat/Supplementary_Information_AIRR_RA/Figure_S2.pdf

http://urr.cat/Supplementary_Information_AIRR_RA/Figure_S3.pdf

http://urr.cat/Supplementary_Information_AIRR_RA/Figure_S4.pdf

http://urr.cat/Supplementary_Information_AIRR_RA/Figure_S5.pdf

http://urr.cat/Supplementary_Information_AIRR_RA/Figure_S6.pdf

http://urr.cat/Supplementary_Information_AIRR_RA/Figure_S7.pdf

http://urr.cat/Supplementary_Information_AIRR_RA/Figure_S8.pdf

http://urr.cat/Supplementary_Information_AIRR_RA/Figure_S9.pdf

### SUPPLEMENTARY TABLES

**TABLE S1. Main clinical and epidemiological characteristics of the study population.** All RA patients, RA patients at baseline, RA patients after 12 weeks of TNFi therapy and control individuals are separately characterized. Abbreviations: N, sample size; NA, not applicable; M, mean; RA, rheumatoid arthritis.

**Supplementary tables can be accessed through the following links:**

http://urr.cat/Supplementary_Information_AIRR_RA/Table_S1.ods

http://urr.cat/Supplementary_Information_AIRR_RA/Table_S2.ods

http://urr.cat/Supplementary_Information_AIRR_RA/Table_S3.ods

http://urr.cat/Supplementary_Information_AIRR_RA/Table_S4.ods

http://urr.cat/Supplementary_Information_AIRR_RA/Table_S5.ods

http://urr.cat/Supplementary_Information_AIRR_RA/Table_S6.ods

http://urr.cat/Supplementary_Information_AIRR_RA/Table_S7.ods

http://urr.cat/Supplementary_Information_AIRR_RA/Table_S8.ods

http://urr.cat/Supplementary_Information_AIRR_RA/Table_S9.ods

http://urr.cat/Supplementary_Information_AIRR_RA/Table_S10.ods

http://urr.cat/Supplementary_Information_AIRR_RA/Table_S11.ods

http://urr.cat/Supplementary_Information_AIRR_RA/Table_S12.ods

http://urr.cat/Supplementary_Information_AIRR_RA/Table_S13.ods

http://urr.cat/Supplementary_Information_AIRR_RA/Table_S14.ods

http://urr.cat/Supplementary_Information_AIRR_RA/Table_S15.ods

http://urr.cat/Supplementary_Information_AIRR_RA/Table_S16.ods

http://urr.cat/Supplementary_Information_AIRR_RA/Table_S17.ods

http://urr.cat/Supplementary_Information_AIRR_RA/Table_S18.ods

http://urr.cat/Supplementary_Information_AIRR_RA/Table_S19.ods

http://urr.cat/Supplementary_Information_AIRR_RA/Table_S20.ods

http://urr.cat/Supplementary_Information_AIRR_RA/Table_S21.ods

http://urr.cat/Supplementary_Information_AIRR_RA/Table_S22.ods

http://urr.cat/Supplementary_Information_AIRR_RA/Table_S23.ods

http://urr.cat/Supplementary_Information_AIRR_RA/Table_S24.ods

http://urr.cat/Supplementary_Information_AIRR_RA/Table_S25.ods

http://urr.cat/Supplementary_Information_AIRR_RA/Table_S26.ods

http://urr.cat/Supplementary_Information_AIRR_RA/Table_S27.ods

http://urr.cat/Supplementary_Information_AIRR_RA/Table_S28.ods

http://urr.cat/Supplementary_Information_AIRR_RA/Table_S29.ods

http://urr.cat/Supplementary_Information_AIRR_RA/Table_S30.ods
